# Plasma metabolomics of primary open-angle glaucoma in three prospective US cohorts and the UK Biobank

**DOI:** 10.1101/2022.02.24.22271483

**Authors:** Oana Zeleznik, Jae H. Kang, Jessica Lasky-Su, A. Heather Eliassen, Lisa Frueh, Clary Clish, Bernard A. Rosner, Tobias Elze, Pirro Hysi, Anthony Khawaja, Janey L. Wiggs, Louis R. Pasquale

**Affiliations:** Channing Division of Network Medicine, Brigham and Women’s Hospital and Harvard Medical School, Boston, Massachusetts; Departments of Nutrition and Epidemiology, Harvard T.H. Chan School of Public Health, Boston, Massachusetts; Broad Institute of Massachusetts Institute of Technology and Harvard, Cambridge, Massachusetts; Department of Biostatistics, Harvard T.H. Chan School of Public Health, Boston, Massachusetts; Department of Ophthalmology, Schepens Research Eye Institute of Massachusetts Eye and Ear, Harvard Medical School, Boston, Massachusetts; Department of Ophthalmology, King’s College London, St. Thomas’ Hospital, London, UK; Department of Twin Research & Genetic Epidemiology, King’s College London, St. Thomas’ Hospital, London, UK; NIHR Biomedical Research Centre at Moorfields Eye Hospital & UCL Institute of Ophthalmology, London, UK; Department of Ophthalmology, Massachusetts Eye and Ear, Harvard Medical School, Boston, Massachusetts; Department of Ophthalmology, Icahn School of Medicine at Mount Sinai, New York, New York

## Abstract

**Purpose:** To better understand the etiologic pathways in glaucoma, we aimed to identify pre-diagnostic plasma metabolites associated with glaucoma risk.

**Methods:** In a case-control study from the Nurses’ Health Study (NHS), NHSII and Health Professionals Follow-Up Study (HPFS), 599 incident primary open-angle glaucoma (POAG) cases (mean time between blood draw and diagnosis was 10.3 years) were 1:1 matched to 599 controls. Plasma metabolites were measured with LC-MS/MS at the Broad Institute (Cambridge, MA, USA); 367 metabolites from 17 metabolite classes passed quality control analyses. For comparison, in a cross-sectional study in the UK Biobank, 168 NMR metabolites (Nightingale, Finland; version 2020) were measured in serum samples from 2,238 prevalent glaucoma cases and 44,723 controls. Metabolites were probit-score transformed for normality; multiple logistic regression was used to identify metabolites associated with POAG in NHS/NHSII/HPFS and glaucoma in UK Biobank. In NHS/NHSII/HPFS, we also used Metabolite Set Enrichment Analysis to identify metabolite classes associated with POAG. All analyses adjusted for established glaucoma risk factors. False discovery rate (FDR) and number of effective tests (NEF) were used to adjust for multiple comparisons.

**Results:** Nine metabolite classes were associated (FDR<0.05) with POAG in NHS/NHSII/HPFS: triglycerides, diglycerides, two lysophospholipids classes [lysophosphatidylcholines and lysophosphatidylethanolamines], and two phospholipid class [phosphatidylethanolamines and phosphatidylcholines] were positively associated, while cholesteryl esters, carnitines, and organic acids and derivatives were inversely associated with POAG risk; further adjustment for covariates minimally altered the results. These associations were particularly stronger for POAG with paracentral visual field loss. In the UK Biobank, notably, triglycerides and phospholipids (from which lysophospholipids are derived through hydrolysis), were confirmed to be associated (p<0.05) with higher glaucoma risk. Also, in the UK Biobank, the metabolites of tyrosine, glucose, and glutamine were positively associated (NEF<0.2) while 3-hydroxybutyrate, acetate, citrate, pyruvate, and lactate (the latter 4 being anionic organic acids) were inversely associated with glaucoma (NEF<0.05).

**Conclusions:** Higher levels of glycerides (diglycerides and triglycerides) and phospholipids were adversely associated with glaucoma in both the NHS/NHSII/HPFS and the UK Biobank, suggesting that they play an important role in glaucoma pathogenesis.

**PRÉCIS:** Higher glyceride and phospholipid levels in pre-diagnostic plasma was associated with glaucoma risk in three cohorts and were associated with prevalent glaucoma in the UK Biobank. Altered lipid metabolism may be etiologically important in glaucoma.

## INTRODUCTION

Glaucoma is a progressive optic neuropathy that is a leading cause of irreversible blindness worldwide.^1^ Primary open-angle glaucoma (POAG) is the most common form, and yet the etiology of this multifactorial disease is poorly understood. Genome-wide association studies have identified >120 genetic loci for POAG,^2^ and the finding of multiple genes in various pathways suggests that there is a complex metabolic network that affects optic nerve health.

The metabolome is the set of small molecule metabolites that are critical for growth and maintenance of cells and tissues,^3, 4^ and represent the end products of environmental factors and gene expression associated with responses to such factors. Metabolomics platforms quantify blood metabolites and can be used to evaluate the etiologic role of metabolic alterations. Multiple proteomic and metabolomics studies of prevalent POAG/glaucoma have been conducted,^5–9^ and some novel markers have been found to be associated with POAG. However, this approach may be problematic for the discovery of changes related to early disease, as consequences of advanced disease or treatment are likely to impact circulating metabolite profiles with the use of prevalent glaucoma cases. In this study, we included 599 incident cases and 599 matched controls in a nested case-control study of pre-diagnostic circulating plasma metabolites from ~10 years before POAG diagnosis, and to confirm the findings, we evaluated the metabolomic data in prevalent cases in the UK Biobank.

## METHODS

### Study Population - Nurses’ Health Study (NHS), NHSII and Health Professional Follow-up Study (HPFS)

We conducted a nested case-control study in the NHS, NHSII and HPFS cohorts from the US. The NHS was initiated in 1976 with 121,700 female registered nurses aged 30–55 years at enrollment; the NHSII began in 1989 with 116,429 female registered nurses aged 25-42 years, and the HPFS was launched in 1986 with 51,529 male health professionals aged 40–75 years. Participants have completed biennial questionnaires that asked about lifestyle and medical conditions, such as glaucoma. POAG cases were identified among participants who self-reported a physician diagnosis of glaucoma on biennial questionnaires. To confirm the self-reports of glaucoma, we asked participants for their consent to obtain relevant medical records from their treating providers. All eye care providers of record were requested to send all available visual fields (VFs) and were mailed a supplementary questionnaire to complete and return. A glaucoma specialist (LRP) reviewed the questionnaire (or medical records sent instead of questionnaires) as well as the VFs in a standardized manner. This questionnaire included items about untreated maximum IOP, any secondary causes of high untreated IOP, filtration angle, structural features of the optic nerve, glaucoma surgery, any VF loss and any secondary conditions that may cause VF loss. Cases we included in analyses had at least two reliable VFs (≤ 20% for false negative rate and false positive rate and ≤ 33% for fixation loss rate) that showed reproducible defects consistent with glaucoma, non-occludable angles in both eyes, and no secondary causes of IOP elevation (e.g., trauma, uveitis, exfoliation syndrome, pigment dispersion syndrome evident on biomicroscopic anterior segment examinations).

Blood samples were collected in 1989–’90 among 32,826 NHS participants, in 1996–’99 among 29,611 NHS2 participants and in 1993–’95 among 18,159 HPFS participants. Participants arranged to have samples drawn and shipped by overnight courier to the laboratory; with sample processing, white blood cell, red blood cell, and plasma aliquots were archived in liquid nitrogen freezers (≤-130^°^C). POAG cases were diagnosed after blood draw until June 1, 2016 (NHS and NHS2), or January 1, 2016 (HPFS). Controls were 1:1 matched to cases on: age, cohort/sex, month and year of blood collection, time of day of blood draw, fasting status (> or ≤8 hours), race/ethnicity, and among women: additional matching on menopausal status and hormone therapy use at blood draw (premenopausal, postmenopausal and on hormone therapy (HT), postmenopausal and not on HT, missing/unknown) and at glaucoma diagnosis. For matching factors and covariates, we used questionnaire data collected as of the blood draw, and if not available, we used biennial questionnaire data prior to the blood sample. The study protocol was approved by the institutional review boards (IRBs) of the Brigham and Women’s Hospital, Harvard T.H. Chan School of Public Health, and Icahn School of Medicine at Mount Sinai. Completion of self-administered questionnaires and returns of blood samples were considered as implied consents by the IRBs. Medical record release consents were obtained for collection of medical records. This research study adhered to the tenets of the Declaration of Helsinki.

### Study Population – UK Biobank

The UK Biobank is an ongoing population-based study initiated in 2006-2010 with over 500,000 participants aged 40-69 years. We used baseline questionnaire as well as metabolomic data (http://www.ukbiobank.ac.uk). The UK Biobank was approved by the National Information Governance Board for Health and Social Care and the National Health Service North West Multicenter Research Ethics Committee (reference number 06/MRE08/65). This research was conducted using the UK Biobank Resource under application number 36741.

At baseline (2006-2010), participants completed a touch screen questionnaire and were considered to have glaucoma if in response to the question, “Has a doctor told you that you have any of the following problems with your eyes?”, they chose glaucoma from the menu. Participants were also considered to have glaucoma if they reported a history of glaucoma surgery or laser on the questionnaire or if they carried an ICD9/10 code for glaucoma (ICD 9: 365.*; ICD10: H40.** (excluding H40.01* and H42.*)).

### Metabolite profiling – NHS, NHSII, HPFS

As described previously,^10–12^ plasma metabolites were profiled using liquid chromatography tandem mass spectrometry (LC-MS) to measure endogenous, polar metabolites and lipids (Broad Institute of MIT and Harvard University (Cambridge, MA)). Among 427 known metabolites, we excluded 60 that were impacted by delayed blood processing,^13^ leaving 367 metabolites for analyses. For metabolites with <10% missing across participant samples, missing values were imputed with 1/2 of the minimum value measured for that metabolite as has been done in prior studies.^14^ All but one included metabolite exhibited good within person reproducibility over a 1-year period, indicating that one blood sample provides a reasonable measure of longer-term exposure (312 out of 313 available 1 year Pearson or intraclass correlations were >0.4).^13^ For 317 (out of 367) metabolites, they were each assigned a metabolite class (30 in total) based on chemical taxonomy; we evaluated 17 metabolite classes which included at least 3 metabolites: steroids and steroid derivatives; carnitines; diglycerides (DGs); triglycerides (TGs); cholesteryl esters; lysophosphatidylethanolamines (LPEs); phosphatidylethanolamines (PEs); lysophosphatidylcholines (LPCs); phosphatidylcholines (PCs); phosphatidylcholine plasmalogens; phosphatidylethanolamines plasmalogens; organoheterocyclic compounds; ceramides; carboxylic acids and derivatives; organic acids and derivatives; nucleosides, nucleotides, and analogues; and sphingomyelins. Metabolite values were transformed to probit scores to scale to the same range and to minimize the influence of skewed distributions.

### Metabolite profiling – UK Biobank

In the UK Biobank, non-fasting baseline plasma samples from a random subset of 119,764 participants (118,466 individuals’ samples collected at baseline and 1298 individuals’ samples collected during repeat-visits) were assessed using targeted high-throughput NMR metabolomics (Nightingale Health Ltd; Helsinki, Finland).^15^ In contrast to LC/MS, in ^1^H NMR spectroscopy, molecules with H atoms yield distinctive spectral shapes with areas under their curves proportional to the molecules’ concentration based on chemical shifts and J coupling splitting patterns determined from quantum mechanics,^16, 17^ thus allowing for detailed quantification. The platform provided quantification of 249 metabolic measures including routine clinical lipids (37 biomarkers in the panel have been certified for diagnostic use), lipoprotein subclasses, fatty acid composition, and several low-molecular weight metabolites such as glycolysis metabolites, ketone bodies and amino acids measured in molar concentration units. Of the 249 measures, we focused on 168 measures that were concentrations of various metabolites (and the 81 remaining measures such as ratios between metabolites, percentages of individual metabolites of total classes or degree of unsaturation were excluded).

### Statistical analysis

#### 1) Model building / covariates - NHS, NHSII, HPFS

Given the matched design, for the individual metabolite analyses and metabolite class analyses, in successive models, we used nested multivariable-adjusted conditional logistic regression models. Model 1 did not adjust for any covariates and evaluated the matched data as is. In Model 2, we adjusted for factors as of blood draw that have a major influence on metabolites and a subset of matching factors where there was imperfect matching between cases and controls (e.g., cohort/sex was perfectly matched, but age was not, so was additionally adjusted for): age (years; matching factor), month of blood draw (matching factor), time of blood draw (matching factor), fasting status (>8 hours or less; matching factor), body mass index (BMI, kg/m^2^), smoking status (never, past, current), and physical activity (metabolic equivalents of task-hours/week). In Model 3, we further added established risk factors for POAG: glaucoma family history, ancestry (non-Hispanic white, black, Asian, Hispanic white), index of socioeconomic status based on 7 census-tract variables and for women: age at menopause (linear age). In Model 4, we added additional POAG risk factors of intake of nitrate,^18^ caffeine,^19^ alcohol,^20^ Alternate Healthy Eating Index excluding alcohol,^21^ and total caloric intake. In Model 5, we further added co-morbidities associated with POAG: hypertension, high cholesterol, diabetes, oral steroid use.

#### 2) Model building / covariates – UK Biobank

As data from the UK Biobank was used to evaluate whether the results from the NHS/NHSII/HPFS are reproduced in an independent population, we used multiple logistic regression to evaluate the association between metabolite levels in relation to prevalent glaucoma. The multiple logistic regression model adjusted for age and age-squared (years; to finely control for age), sex, ethnicity (Caucasian, Black and other), smoking status (never, past and current smoker), number of cigarettes smoked daily among current smokers, alcohol intake frequency (daily or almost daily, 3-4 times a week, 1-2 times a week, 1-3 times a month, special occasions only, never), coffee and tea consumption (cups per day), physical activity (Metabolic Equivalent of Task (MET)-hours/week), Townsend deprivation index (range: −6 to 11; a higher index score indicates more relative poverty for a given residential area), BMI (kg/m^2^), systolic blood pressure (mm Hg), history of diabetes (yes or no), history of cardiovascular disease, systemic beta blocker use, use of statin drugs and spherical equivalent. We included 46,961 participants (2238 glaucoma cases and 44,723 controls) with complete data on metabolites and covariates.

#### 3) Analytic approach – all cohorts

All analyses were performed with SAS 9.4 and R 3.4.1. For analyses of individual metabolites, metabolite values were used as continuous variables (per 1 standard deviation (SD) increase) to calculate linear trend p-values. We estimated the odds ratios (OR) and 95% confidence intervals (CIs) per 1 SD increase in metabolite levels.

For evaluating individual metabolites, “number of effective tests” (NEF)^22^ was used to adjust for multiple comparisons as NEF has the advantage of accounting for the high correlation structure of the metabolomics data. NEF-p<0.05 was considered statistically significant and NEF-p<0.2 was considered worthy of additional analysis given that this was an exploratory study.

For NHS/NHSII/HPFS, for metabolite class analyses, Metabolite Set Enrichment Analysis was used. As metabolite classes overall are not correlated, to adjust for multiple comparisons, False Discovery Rate (FDR)^23^ was used. FDR<0.05 was considered statistically significant, while FDR<0.2 was considered nominally significant and worth considering in future analyses given the hypothesis-generating aspect of this study.

## RESULTS

### Study population

Among 599 cases, 74.3% were female, with a mean (SD) age at blood draw of 58.0 (SD=8.0) years and at diagnosis of 68.3 (SD=9.2) years. Mean time between blood draw to diagnosis was 10.3 years. Controls were similar to cases for the matching factors. Distributions of POAG risk factors were generally in the expected directions for cases and controls (**Table 1**).

**Table 1.** **Participant characteristics in the Nurses’ Health Study, Nurses’ Health Study II and Health Professionals Follow-up Study at the time of blood collection\***

|  | **Cases** | **Controls** |
| --- | --- | --- |
|  | **n=599** | **n=599** |
| **Age, years<sup>†</sup>** | 58.0 (8.0) | 57.9 (7.9) |
| **Men, n (%)<sup>†</sup>** | 154 (25.7) | 154 (25.7) |
| **Race/ethnicity, n (%)<sup>†</sup>** |  |  |
| Non-Hispanic Caucasian | 577 (96.3) | 591 (98.7) |
| African | 8 (1.3) | 3 (0.5) |
| Asian | 6 (1.0) | 1 (0.2) |
| Hispanic | 8 (1.3) | 4 (0.7) |
| **Time of blood draw, n (%)<sup>†</sup>** |  |  |
| 12:00am-7:59am | 70(12.2) | 74 (12.7) |
| 8:00am-9:59am | 293 (51.2) | 321 (55.2) |
| 10:00am-11:59am | 124 (21.7) | 121 (20.8) |
| 12:00pm-11:59pm | 85 (14.9) | 65 (11.2) |
| **Season of blood draw, n (%)<sup>†</sup>** |  |  |
| Winter | 137 (22.9) | 126 (21.0) |
| Spring | 223 (37.2) | 206 (34.4) |
| Summer | 202 (33.7) | 218 (36.4) |
| Autumn | 37 (6.2) | 49 (8.2) |
| **Smoking status, n (%)** |  |  |
| Never | 305 (51.3) | 301 (50.8) |
| Past | 244 (41.0) | 248 (41.8) |
| Current | 46 (7.7) | 44 (7.4) |
| **Body mass index, kg/m<sup>2</sup>** | 25.4 (4.7) | 25.5 (4.2) |
| **Physical activity, MET-hours/week** | 23.2 (34.5) | 22.4 (29.9) |
| **Family history of glaucoma, n (%)** | 180 (31.1) | 81 (14.4) |
| **Age at menopause, years** | 49.5 (3.6) | 49.4 (3.4) |
| **Socio-economic status index score** | 0.0 (4.9) | 0.0 (4.8) |
| **Nitrate intake, mg/day** | 152.3 (88.0) | 157.7 (83.1) |
| **Alcohol intake, g/day** | 7.1 (11.2) | 6.9 (12.0) |
| **Caffeine intake, mg/day** | 263.1 (233.6) | 261.8 (232.5) |
| **Alternative Healthy Eating Index score** | 48.3 (11.0) | 48.7 (11.0) |
| **Total caloric intake, kcal/day** | 1841.0 (539.5) | 1832.7 (550.0) |
| **Hypertension, n (%)** | 153 (25.5) | 141 (23.5) |
| **High cholesterol, n (%)** | 193 (32.2) | 188 (31.4) |
| **Diabetes, n (%)** | 24 (4.0) | 19 (3.2) |

Abbreviation: MET=metabolic equivalent of task
\*Values are means ± standard deviation or numbers (percentages), are standardized to the age distribution of the study population and based on those with non-missing values.
<sup>†</sup>Cases and controls were 1:1 matched based on age (all >40 years), cohort/sex, month and year of blood collection, time of day of blood draw, fasting status (> or ≤8 hours), race/ethnicity, and among women: additional matching on menopausal status and hormone therapy use at blood draw (premenopausal, postmenopausal and on hormone therapy (HT), postmenopausal and not on HT, missing/unknown) and at glaucoma diagnosis. All cases and controls reported eye exams in the index period of the matched cases' diagnosis date.
<sup>‡</sup>Alternative Healthy Eating Index scores rang from 0 to 100; a higher score reflects a healthier diet.

### Relation between individual metabolites and POAG

**Supplementary Table S1** presents the individual associations between the 367 metabolites and POAG; 6 metabolites that were at least nominally significant at p<0.05 in any of the models (Model 1 through Model 5) are plotted in **Figure 1**. In Model 1 (model incorporating matching) results, all 6 metabolites were nominally significant, including two diglycerides (DG(36:2), DG(34:1)), one triglyceride (TG(52:2)), two LPCs (LPC(16:0) and Na_LPC(16:0)) and one LPE(16:0); of these, LPE(16:0), TG(36:2), TG(52:2) and Na-LPC(16:0) were NEF-p<0.2. In successive models, the associations were generally similar, although the significance was attenuated with additional adjustment for covariates. In Model 2, where we more finely adjusted for matching factors (e.g., fasting status, time of day) and other major determinants of variability in metabolites, such as BMI, only the glycerides were NEF-p<0.2. However, in successive models of Model 3 and 4 (addition of POAG established and suspected risk factors), and Model 5 (further adjust for co-morbidities), we observed that while none of the metabolites were NEF<0.05, all were nominally significant and the direction of associations for the metabolites was similar.

**Figure 1.**
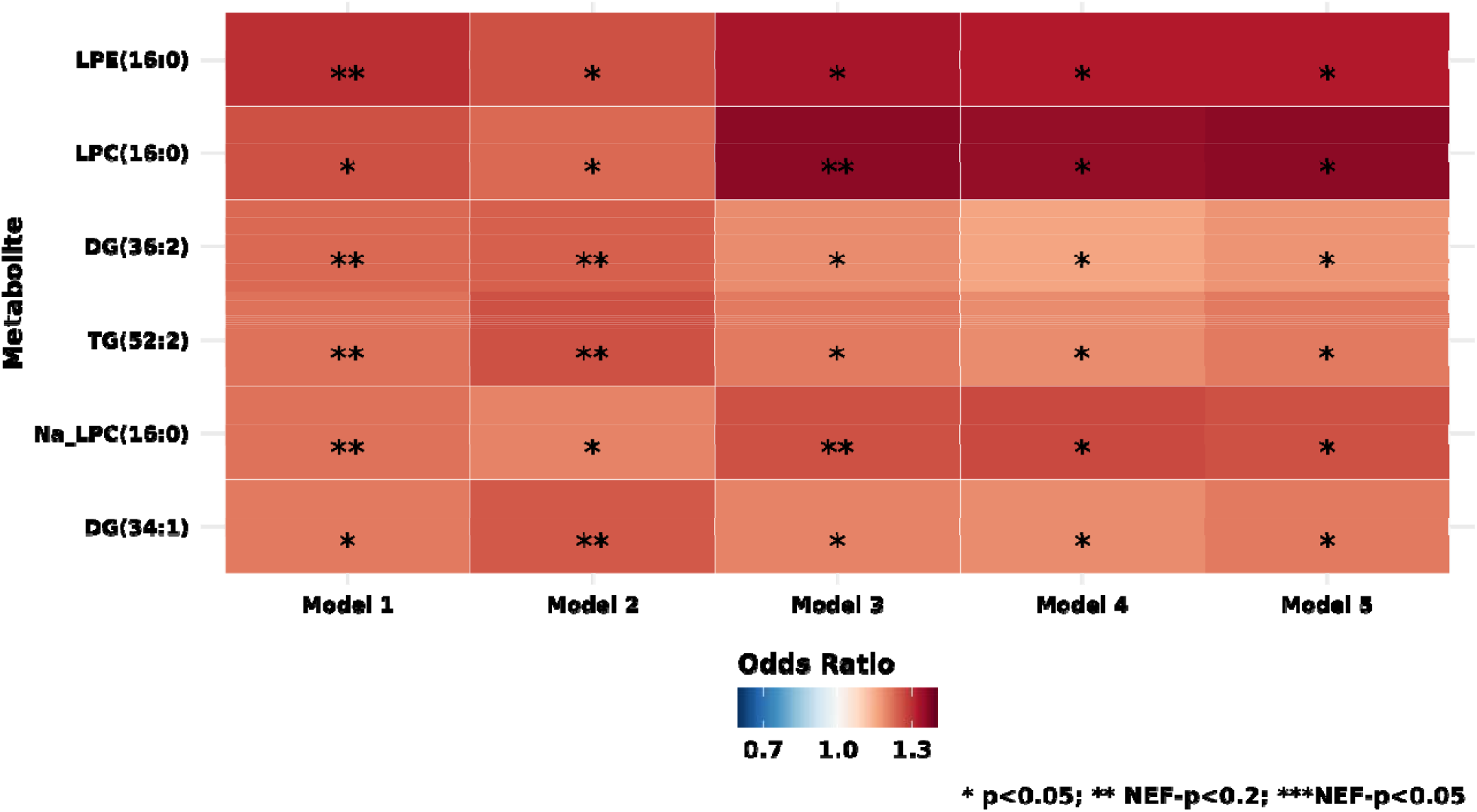
Individual metabolites among the n=367 metabolites evaluated that were significant across the various nested multiple conditional logistic regression models of primary open-angle glaucoma (599 cases and 599 controls). **Model 1**: basic model, adjusting for matching factors only (see Table 1); **Model 2** (factors that affect metabolite levels and matching factors as matching was imperfect): Model 1 + age, sex, smoking status, BMI, physical activity, time of day of blood draw, month of blood draw, fasting status; **Model 3** (established risk factors for primary open-angle glaucoma (POAG)): Model 2 + family history of glaucoma, socioeconomic index based on census tract data, race/ethnicity and age at menopause; **Model 4** (potential modifiable dietary risk factors for POAG): Model 3 + nitrate intake, caffeine intake, alcohol intake, caloric intake; **Model 5** (systemic comorbidities / drugs suggested to be associated with POAG in some studies): Model 4 + hypertension, high cholesterol, diabetes and oral steroid use. * p<0.05; ** Number of effective tests (NEF)<0.2; *** NEF<0.05

### Relation between metabolite classes and POAG

**Figure 2** shows the results of evaluating metabolite classes and POAG; results for all 17 metabolite classes are plotted. Model 1 showed 9 classes that were associated: 6 lipid metabolite classes of triglycerides, LPCs, diglycerides, LPEs, PCs, and PEs were FDR-significantly associated with POAG risk, while cholesteryl esters, carnitines and organic acids and derivatives (which includes amino acids) were inversely associated with POAG risk. In Model 2, results were similar, although among lipids, for phosphatidylethanolamines, the FDR was ≥0.2. Similarly, Models 3, 4 and 5, with the addition of POAG risk factors in models, the adverse associations with LPCs, LPEs, and diglycerides (in Model 5) and the inverse associations with cholesteryl esters and organic acids and derivatives (which includes amino acids) were robust and FDR-significant. For carnitines and phosphatidylethanolamines, the FDR was ≥0.2 and for PCs and triglycerides, the FDR was <0.2; interestingly, in Model 5, an inverse association with sphingomyelins was observed at FDR<0.2.

### Relation between metabolite classes and POAG subtypes defined by visual field loss patterns

POAG is multifactorial and clinically heterogeneous. Investigating heterogeneity in VF loss patterns for POAG may provide new etiologic insights as different types of optic nerve damage manifest as distinct VF loss patterns. For example, glaucomatous paracentral scotomas have been associated with more systemic risk factors compared to peripheral VF loss.^24–26^ Therefore, we separately evaluated the associations between metabolite classes and POAG defined by VF loss patterns (paracentral (**Figure 3a;** n=178 cases) versus peripheral VF loss (**Figure 3b;** n=331 cases)); of the 599 cases, VF loss pattern derived from Humphrey visual field test were available in 509 cases. As shown in Figure 3a and Figure 3b, overall, more metabolite classes (9 classes versus 5 classes) were significantly associated with POAG with paracentral VF loss versus peripheral VF loss. In addition to the three classes associated with peripheral VF loss - namely diglycerides, LPCs, and LPEs - for POAG with paracentral VF loss, the classes of triglycerides, PEs, and PCs were also adversely associated at FDR<0.05. In addition to cholesteryl esters and organic acids and derivatives being inversely associated with POAG with peripheral VF loss, the class of carnitines was also inversely associated with POAG with paracentral VF loss at FDR<0.05.

**Figure 2.**
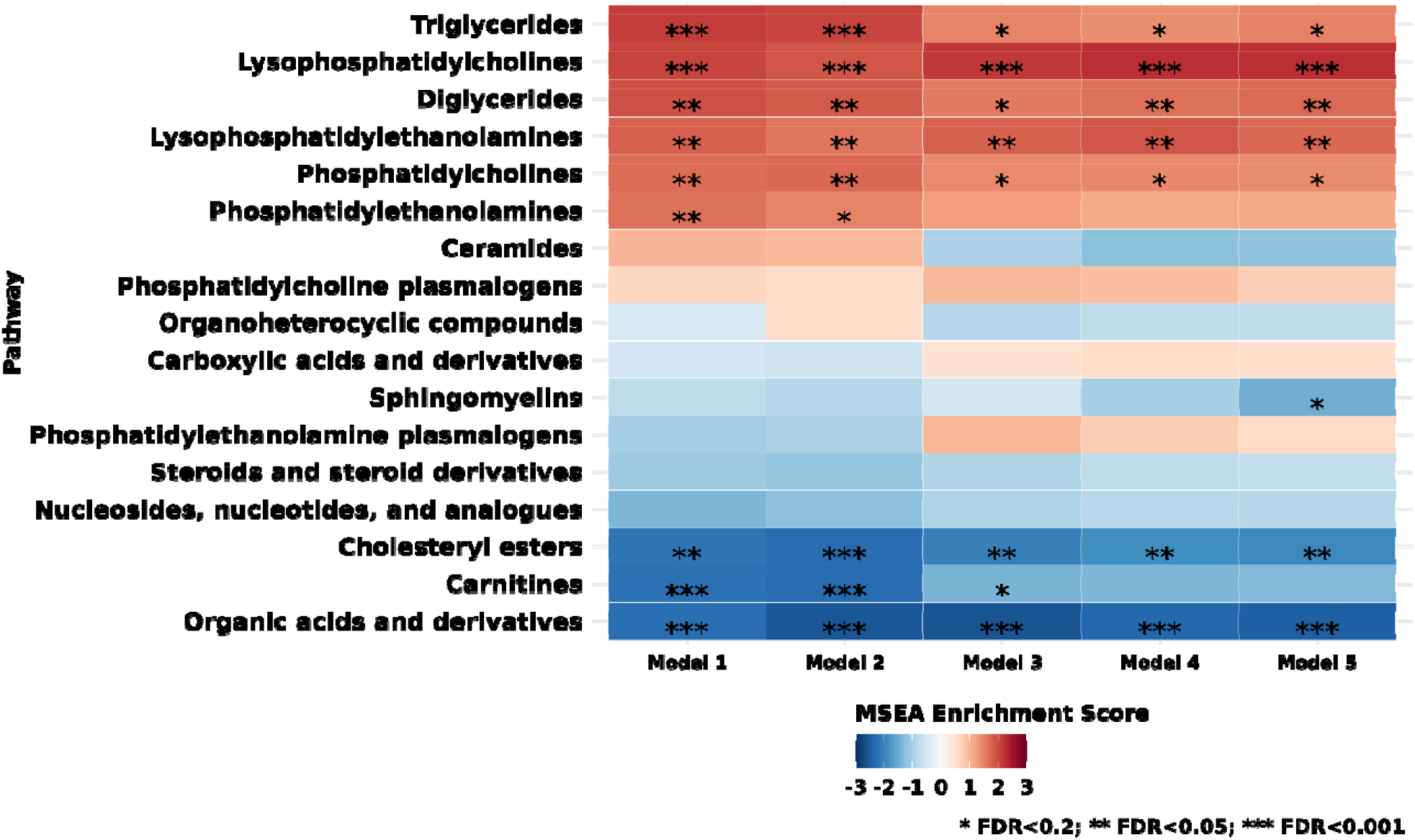
Metabolite classes (n=17) evaluated in various nested multiple conditional logistic regression models of primary open-angle glaucoma (POAG; 599 cases and 599 controls). **Model 1**: basic model, adjusting for matching factors only (see Table 1); **Model 2** (factors that affect metabolite levels and matching factors as matching was imperfect): Model 1 + age, sex, smoking status, BMI, physical activity, time of day of blood draw, month of blood draw, fasting status; **Model 3** (established risk factors for primary open-angle glaucoma (POAG)): Model 2 + family history of glaucoma, socioeconomic index based on census tract data, race/ethnicity and age at menopause; **Model 4** (potential modifiable dietary risk factors for POAG): Model 3 + nitrate intake, caffeine intake, alcohol intake, caloric intake; **Model 5** (systemic comorbidities / drugs suggested to be associated with POAG in some studies): Model 4 + hypertension, high cholesterol, diabetes and oral steroid use. * False Discovery Rate (FDR)<0.2; ** FDR<0.05; *** FDR<0.001

**Figure 3.**
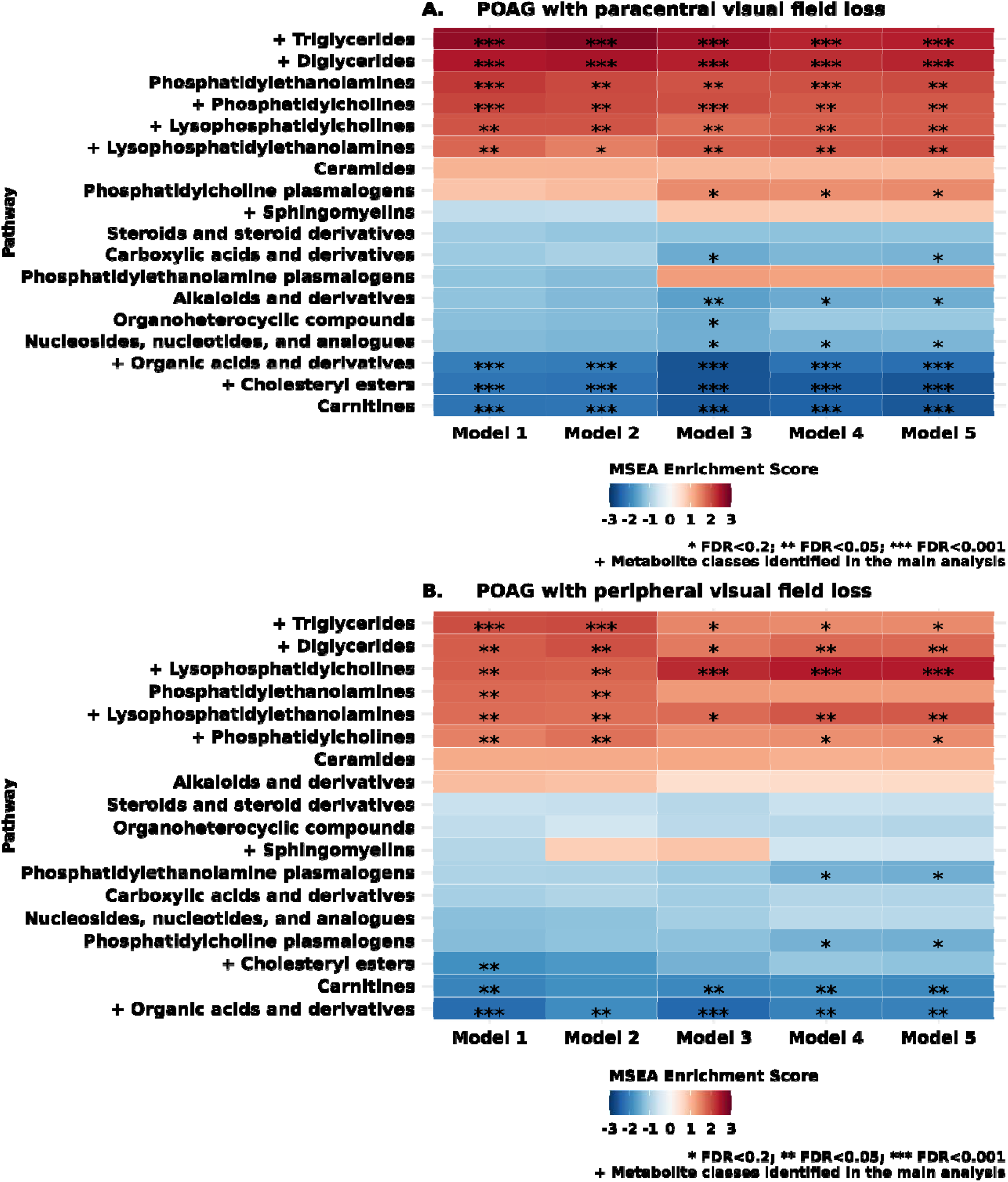
Metabolite classes (n=17) evaluated in various nested multiple logistic regression models of POAG with paracentral visual field loss (178 cases and 599 controls) in panel A and POAG with peripheral visual field loss (331 cases and 599 controls) in panel B, in NHS/NHSII/HPFS. Metabolite classes are plotted in the same as in the main analysis (Figure 2). **Model 1**: basic model, adjusting for matching factors only; **Model 2** (factors that influence metabolites): age + smoking status + BMI + physical activity + time of day (as matching imperfect) + month of blood draw (season, as matching imperfect); **Model 3** (established risk factors for POAG): M2 + family history of POAG + SES + race + age at menopause; **Model 4** (established risk factors for POAG): M3 + nitrate intake + caffeine intake + alcohol intake + alternate healthy eating index + caloric intake; **Model 5** (co-morbidities / drugs that have been associated with POAG): M4 + hypertension + high cholesterol + diabetes + oral/inhaled steroid use. * False Discover Rate (FDR)<0.2; ** FDR<0.05; *** FDR<0.001

### Secondary analyses

Because prior studies of triglycerides in relation to health outcomes^27, 28^ have observed differing associations by the degree of saturation and number of carbon atoms, in exploratory analyses, we further plotted the associations as a function of the number of double bonds in triglycerides and the number of carbon atoms. Interestingly, as found with diabetes and cardiovascular disease,^27, 28^ in general, those triglycerides with fewer carbon atoms and double bonds were adversely associated with POAG risk while those with higher carbon atoms and double bonds were inversely associated with POAG risk (Figure 4).

**Figure 4.**
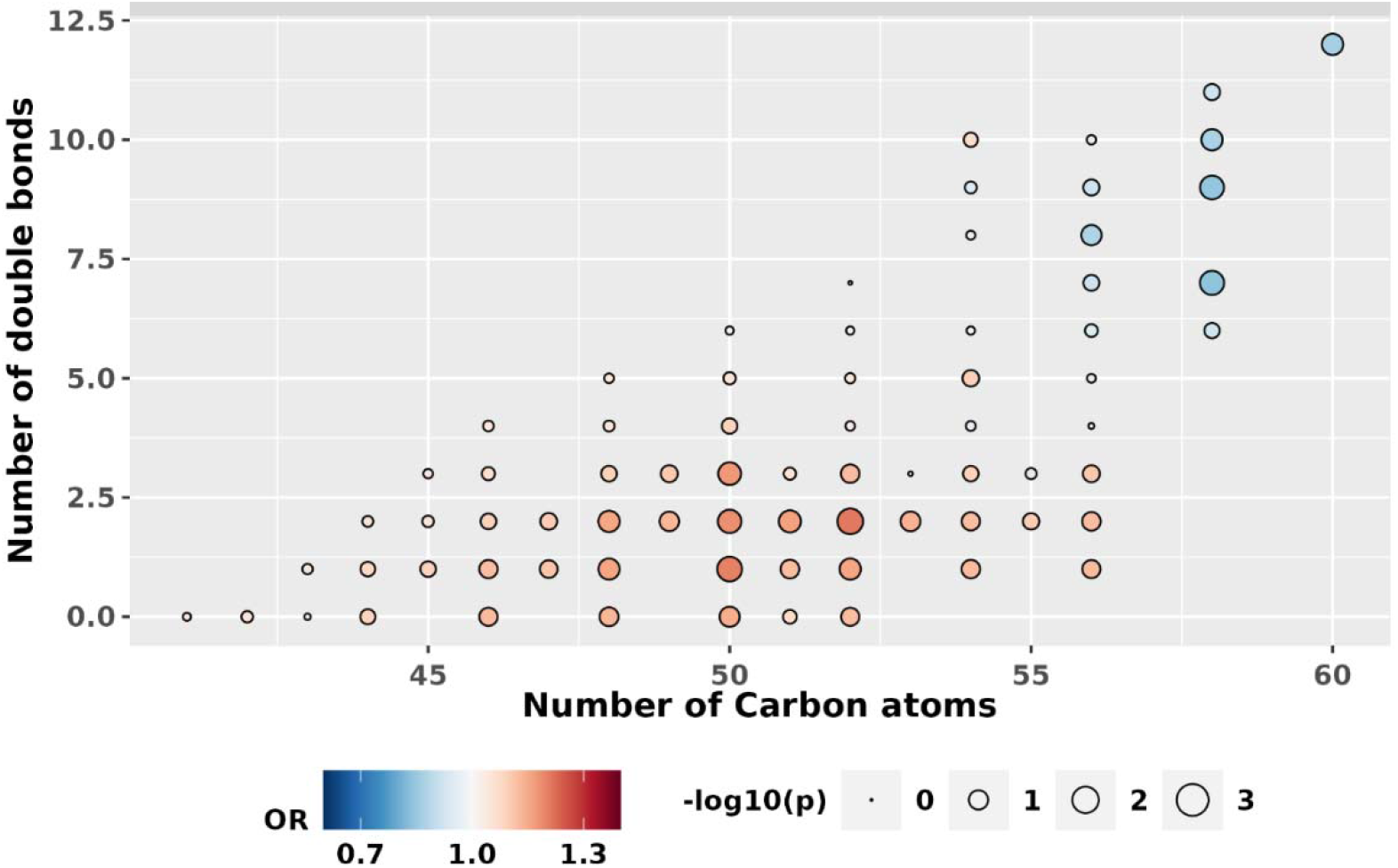
POAG association pattern among triglycerides (TGs) in NHS/NHSII/HPFS (599 cases and 599 controls). Results are shown by the number of carbon atoms (fatty acyl chain length) and double bonds (saturation) of each triglyceride. Each triglyceride is represented by a circle. The color of the circle correlates with the association direction (red: positive, blue: inverse) and the size of the circle corelates with the statistical significance (larger circles represent smaller p-values).

To evaluate whether associations differed by subgroups, in exploratory analyses, we assessed whether results varied by age (**Supplementary Figure S1**), sex (**Supplementary Figure S2**), BMI (**Supplementary Figure S3**), time to diagnosis (**Supplementary Figure S4**), and history of glaucoma (**Supplementary Figure S5**). Although no major differences were observed in various subgroups, there were some suggestions that the associations were strongest in those with BMI>25 kg/m^2^ and those diagnosed within the decade after blood draw.

### UK Biobank results

To assess whether the associations observed in NHS/NHS2/HPFS might also be observed in the UK Biobank, we conducted metabolomic analyses of the outcome of glaucoma, defined based on self-reported glaucoma, use of glaucoma medications and ICD codes (2238 glaucoma cases and 44723 non-cases). In general, glaucoma cases were older, had more diabetes, and higher systolic blood pressure (**Table 2**).

**Table 2.** UK Biobank participant characteristics at time of blood draw*.

|  | **Glaucoma**<br>**Cases**<br>**n=2238** | **Controls**<br>**n=44723** |
| --- | --- | --- |
| **Age, years** | 61.2 (6.4) | 56.8 (8.0) |
| **Men, n (%)** | 1178 (52.6) | 20369 (45.5) |
| **Smoking status, n (%)** |  |  |
| Never | 1177 (52.6) | 24891 (55.7) |
| Past | 853 (38.1) | 15315 (34.2) |
| Current | 208 (9.3) | 4517 (10.1) |
| **Physical activity, MET-hours/week** | 2508.8 (2483.6) | 2463.6 (2429.9) |
| **Body mass index, kg/m<sup>2</sup>** | 27.8 (4.5) | 27.3 (4.5) |
| **Race/Ethnicity, n (%)** |  |  |
| White | 2049 (91.6) | 41339 (92.4) |
| Black | 64 (2.9) | 1272 (2.8) |
| Asian | 76 (3.4) | 1078 (2.4) |
| Other | 49 (2.2) | 1034 (2.3) |
| **Coffee consumption, cups/day** | 1.9 (1.7) | 1.9 (1.8) |
| **Tea consumption, cups/day** | 3.1 (2.0) | 3.1 (2.1) |
| **Alcohol intake, n (%)** |  |  |
| Never or special occasion only | 543 (24.3) | 11213 (25.1) |
| **Systolic blood pressure, mm Hg** | 141.2 (17.9) | 137.3 (18.1) |
| **Diabetes, n (%)** | 256 (11.4) | 3001 (6.7) |
| **Coronary artery disease, n (%)** | 210 (9.4) | 2342 (5.2) |
| **Cholesterol, mmol/l** | 5.6 (1.2) | 5.7 (1.1) |

\*Values are means (standard deviations) or numbers (percentages)

In multivariable-adjusted analyses of individual metabolites (**Figure 5; Supplementary Table S2**), we observed that 8 triglyceride metabolites as well as phospholipids (from which lysophospholipids are derived through hydrolysis) were nominally associated with higher glaucoma risk; tyrosine (NEF<0.05) and glucose (NEF<0.05) were significantly associated with higher glaucoma risk, while specific organic acids and derivatives, such as acetate, 2-hydroxybutyrate, citrate, pyruvate and lactate were inversely associated at NEF<0.05. (In NHS/NHS2/HPFS (**Supplementary Table S1**), tyrosine and valine were included as carboxylic acids and derivatives; glutamine and phenylalanine were included as organic acids and derivatives; however, glucose, acetate, 3-hydroxybutyrate, citrate, pyruvate and lactate data were not available).

**Figure 5.**
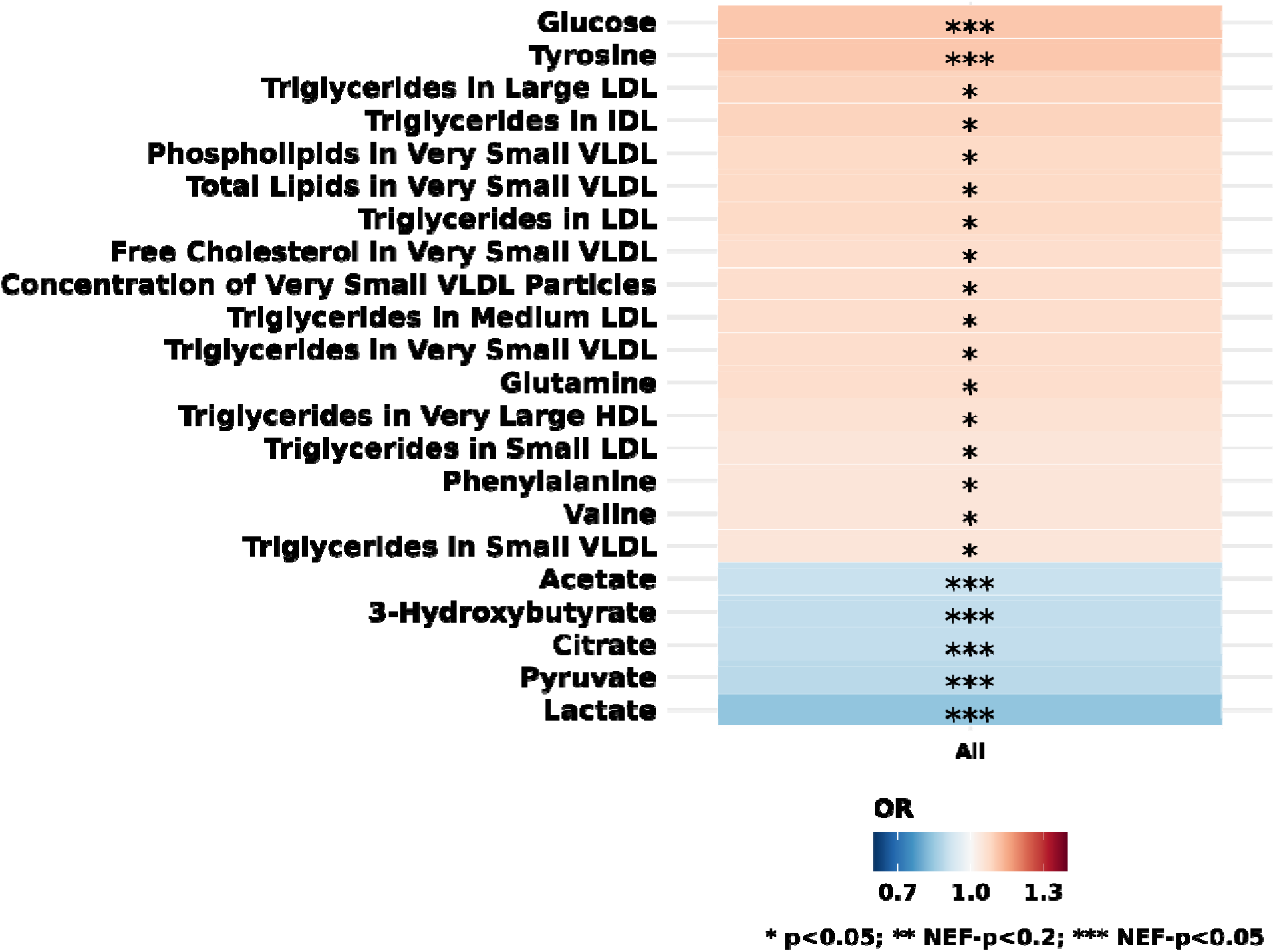
Individual metabolites (out of N=168) that were nominally significantly associated with prevalent POAG in the UK Biobank (2238 cases, 44723 controls). Multiple logistic regression model includes age, sex, smoking status, physical activity, BMI, ethnicity, spherical equivalent, coffee consumption, tea consumption, alcohol intake, systolic blood pressure, cholesterol level, diabetes, coronary artery disease, and statin use. * p<0.05; ** Number of effective tests corrected p-value (NEF)<0.2; *** NEF<0.05

## DISCUSSION

In this nested case-control study of pre-diagnostic plasma metabolites in relation to POAG (n=1198), with a mean 10.3 years between blood draw and diagnosis, we observed that higher levels of diglycerides, triglycerides and phospholipids were adversely associated with POAG in the NHS/NHSII/HPFS, with stronger associations for POAG with paracentral VF loss. The adverse associations with glycerides and phospholipids in relation to prevalent glaucoma were confirmed in the cross-sectional UK Biobank, underscoring their importance in the etiology of glaucoma. While this study was the first to evaluate the relation between pre-diagnostic plasma metabolites, the replication of our findings in a prevalent glaucoma dataset do support a role for altered lipid regulation in glaucoma.

A systematic review^9^ has identified 13 studies to date on the metabolomics of open-angle glaucoma. Of these, 3 evaluated serum,^29–31^ and 3 evaluated plasma^32–34^ while others have evaluated aqueous humor,^35–38^ tear^39^ and optic nerve^40^ samples; these studies have collectively assessed ~140 different metabolites. Compared to existing studies (where the largest study included 211 cases and 295 controls^30^), our study was unique in that the sample size in NHS/NHS2/HPFS was the largest to date (599 cases and 599 controls), did not use convenience sample of controls (e.g., those with cataract or other non-glaucoma eye conditions) and importantly, evaluated pre-diagnostic plasma collected a mean of 10.3 years before diagnosis for POAG cases versus evaluating blood samples from prevalent or newly diagnosed cases, which may be affected by disease or treatment. In addition, our study evaluated metabolomic and glaucoma data from the UK Biobank, to provide independent confirmation of findings, although the metabolomics platforms did not overlap substantially. Nonetheless, the nominal replication of adverse associations between selected lipid species with glaucoma is remarkable as the assay systems were fundamentally different: LC-MS in NHS/NHS2/HPFS vs NMR spectroscopy in UK Biobank. Of note, the systematic review identified 12 metabolites that have been identified across the various metabolomics studies as being associated with open-angle glaucoma in at least 2 studies; of these, two were phospholipids (PCs), 2 were organic acids and derivatives (including amino acids), and 2 were carnitines.^9^ Our results are consistent with prior studies in identifying phospholipids and various organic acids and derivatives in both the NHS/NHS2/HPFS and UK Biobank, while also showing support for the role of lipids, particularly diglycerides, triglycerides and other lipids (e.g., cholesteryl esters).

### Triglycerides and cholesteryl esters

The relation between triglycerides and glaucoma has been conflicting, with studies showing an adverse association,^41^ inverse association^42^ and null association (Mendelian randomization study^43^). A systematic review^44^ of 12 studies reported a significantly higher risk of open-angle glaucoma with hyperlipidemia as well as hypertriglyceridemia (OR=1.42; 95%CI 1.04, 1.93; based on pooling of 4 studies). Our study confirmed this adverse association between higher triglycerides and POAG risk in two populations. This finding is consistent with the discovery of POAG loci near genes related to cholesterol metabolism (*ABCA1* and *CAV1/2)*. One biological mechanism for this association may be intraocular pressure-related: hypertriglyceridemia can lead to increased blood viscosity, which causes elevation of episcleral venous pressure^45, 46^ and higher IOP. Indeed, a review^44^ reported that higher triglyceride level was significantly associated with modest increases of IOP. Interestingly, we observed that the relationship with triglycerides were complex. Triglycerides that are shorter and more saturated were adversely associated while those that are longer and are more unsaturated were inversely associated with POAG, possibly because such triglycerides may be markers of a healthier cardiometabolic profile.^28^ Finally, triglycerides showed stronger associations with POAG with paracentral VF loss than that with peripheral loss, further underscoring the observation that this subtype is more strongly associated with systemic factors. As for cholesteryl esters, there is limited data in relation to glaucoma; however, it is known that the ratio of cholesterol to esterified cholesterol is altered with age and in several neurodegenerative diseases.^47^

### Phospholipids

We observed adverse associations between several phospholipid classes and POAG risk. PCs/PEs and their partial hydrolysis products, LPCs/LPEs, are implicated in aging^48–50^ as well as type 2 diabetes^51^, and play key roles in mitochondrial dynamics.^52^ PEs and PCs are abundant membrane phospholipids that provide membrane structure and facilitate vacuolar delivery and cytokinesis. However, LPCs are pro-inflammatory and pro-oxidant,^53^ and they may play a role in the autotaxin-lysophophatidic acid pathway in intraocular pressure regulation.^35, 37, 54–57^ Our results are consistent with other targeted lipidomic studies of glaucoma that have revealed higher specific phospholipids in blood,^9^ the trabecular meshwork or aqueous humor of glaucomatous eyes.^55, 56, 58, 59^

### Mitochondrial dysfunction, carnitines and organic acids and derivatives

Mitochondrial dysfunction has been hypothesized as a key component of POAG pathophysiology.^60–66^ Plasma metabolomic markers of mitochondrial function may include acylcarnitines, which transport acyl-groups (organic acids and fatty acids) into the mitochondria so that they can be broken down to produce energy in a process known as beta-oxidation. In our study, POAG cases had lower levels of acylcarnitines, particularly the long-chain acylcarnitines. Interestingly, high-fructose diets were associated with a decrease in long-chain acylcarnitines, suggesting a decrease in mitochondrial *β*-oxidation and an increase in lipid peroxidation, which also led to higher triglycerides.^67^ Carnitine has also been shown to be neuroprotective in a glaucoma animal model.^68^ This is consistent with several cross-sectional studies of glaucoma that have also identified alterations in the carnitine pool.^33, 35, 69^

Related to the importance of mitochondrial health, greater use as an energy source of ketone bodies, such as higher 3-hydroxybutyrate versus glucose may also be important. In the UK Biobank, a higher level of 3-hydroxybutyrate was inversely associated with POAG risk. Ketone bodies have been associated with greater mitochondrial efficiency and have neuroprotective properties.^70^ This is consistent with a large genetic pathway analysis study that identified the butanoate pathway, which is involved in generating precursors for ketone bodies, as being important for POAG etiology, ^71^ a study that observed suggestive lower risk of POAG with paracentral VF loss with a plant-based low carbohydrate diet^72^ and another study analyzing genes encoding mitochondrial proteins that implicated lipid and carbohydrate metabolism as being important in POAG.^73^

Among the most inversely associated organic acids and derivatives were N1-Acetylspermidine and N1,N12-Diacetylspermine (Supplementary Table S1). Spermine and spermidine are amines that also regulate mitochondrial membrane potential and have neuroprotective effects;^33^ these metabolites have also been inversely associated with POAG in the study by Leruez et al.^33^ Tyrosine is another that has been identified by Leruez et al.^33^ and was found to be associated with glaucoma in the UK Biobank and thus warrants further study.

A limitation of our data is that our NHS/NHS2/HPFS study population was relatively homogeneous, with mostly White health professionals; therefore, our findings may not be generalizable to other populations with different race and ethnicity composition. Additionally, there may have been residual confounding by other unmeasured factors. A further potential limitation is that our blood samples were collected at one timepoint; However, we included only those metabolites with good correlations for within person stability over at least 1 year. In addition, the UK Biobank data was cross-sectional and included prevalent cases of glaucoma, who would have been comprised mostly of POAG cases, but may have also included those with other types of glaucoma, which would have led to biases due to disease misclassification. Also, there was the possibility in the UK Biobank that metabolite levels may have been influenced by glaucoma treatment; thus, our results need to be interpreted cautiously. However, the confirmation of the associations with triglycerides and phospholipids provided support to the prospective associations observed in NHS/NHS2/HPFS.

Our study had several strengths. To our knowledge, our study in NHS/NHS2/HPFS is the first study assessing the associations of pre-diagnostic metabolites collected ~ 10 years before diagnosis and POAG risk, with a relatively large sample size of 599 cases and 599 matched controls. Additional strengths include the detailed covariate information and long time between blood draw and diagnosis date (mean=10.3 years) and the availability of the UK Biobank data to confirm findings.

Overall, these results provide new insights into the etiology of POAG. Our data implicate dysregulation in lipid metabolism and mitochondrial function in glaucoma etiology and suggest new targets for glaucoma prevention or therapies.

## Data Availability

Information including the procedures to obtain and access data from the Nurses Health Studies and Health Professionals Follow-up Study is described at https://www.nurseshealthstudy.org/researchers (contact) and https://sites.sph.harvard.edu/hpfs/for-collaborators/

## ACKNOWLEDGMENTS

This work was supported by grants from the National Institutes of Health: R01 EY015473 (LRP), NCI UM1 CA186107, U01 CA167552, U01 CA176726, R01 CA49449, R01 CA67262 and an unrestricted challenge grant to Icahn School of Medicine at Mount Sinai, Department of Ophthalmology from Research to Prevent Blindness (LRP). The content is solely the responsibility of the authors and does not necessarily represent the official views of the National Institutes of Health. APK is supported by a UK Research and Innovation Future Leaders Fellowship (Medical Research Council MR/T040912/1).

## References

1. Quigley HA. Number of people with glaucoma worldwide. Br J Ophthalmol. 1996;80:389–393.

2. Gharahkhani P, Jorgenson E, Hysi P, et al. Genome-wide meta-analysis identifies 127 open-angle glaucoma loci with consistent effect across ancestries. Nat Commun. 2021;12:1258.

3. Goodacre R, Vaidyanathan S, Dunn WB, et al. Metabolomics by numbers: acquiring and understanding global metabolite data. Trends Biotechnol. 2004;22:245–252.

4. Roberts LD, Souza AL, Gerszten RE, Clish CB. Targeted metabolomics. Curr Protoc Mol Biol. 2012;Chapter 30:Unit 30 32 31–24.

5. Roedl JB, Bleich S, Schlotzer-Schrehardt U, et al. Increased homocysteine levels in tear fluid of patients with primary open-angle glaucoma. Ophthalmic Res. 2008;40:249–256.

6. Pieragostino D, Agnifili L, Fasanella V, et al. Shotgun proteomics reveals specific modulated protein patterns in tears of patients with primary open angle glaucoma naive to therapy. Mol Biosyst. 2013;9:1108–1116.

7. Pieragostino D, Bucci S, Agnifili L, et al. Differential protein expression in tears of patients with primary open angle and pseudoexfoliative glaucoma. Mol Biosyst. 2012;8:1017–1028.

8. Ghaffariyeh A, Honarpisheh N, Shakiba Y, et al. Brain-derived neurotrophic factor in patients with normal-tension glaucoma. Optometry. 2009;80:635–638.

9. Wang Y, Hou XW, Liang G, Pan CW. Metabolomics in Glaucoma: A Systematic Review. Invest Ophthalmol Vis Sci. 2021;62:9.

10. Vardhan SA, Haripriya A, Ratukondla B, et al. Association of Pseudoexfoliation With Systemic Vascular Diseases in a South Indian Population. JAMA Ophthalmol. 2017;135:348–354.

11. O’Sullivan JF, Morningstar JE, Yang Q, et al. Dimethylguanidino valeric acid is a marker of liver fat and predicts diabetes. J Clin Invest. 2017;127:4394–4402.

12. Mascanfroni ID, Takenaka MC, Yeste A, et al. Metabolic control of type 1 regulatory T cell differentiation by AHR and HIF1-alpha. Nat Med. 2015;21:638–646.

13. Townsend MK, Clish CB, Kraft P, et al. Reproducibility of metabolomic profiles among men and women in 2 large cohort studies. Clin Chem. 2013;59:1657–1667.

14. Zeleznik OA, Eliassen AH, Kraft P, et al. A Prospective Analysis of Circulating Plasma Metabolites Associated with Ovarian Cancer Risk. Cancer Res. 2020;80:1357–1367.

15. Julkunen H, Cichonska A, Slagboom PE, et al. Metabolic biomarker profiling for identification of susceptibility to severe pneumonia and COVID-19 in the general population. Elife. 2021;10.

16. Soininen P, Kangas AJ, Wurtz P, et al. Quantitative serum nuclear magnetic resonance metabolomics in cardiovascular epidemiology and genetics. Circ Cardiovasc Genet. 2015;8:192–206.

17. Mihaleva VV, Korhonen SP, van Duynhoven J, et al. Automated quantum mechanical total line shape fitting model for quantitative NMR-based profiling of human serum metabolites. Anal Bioanal Chem. 2014;406:3091–3102.

18. Kang JH, Willett WC, Rosner BA, et al. Association of Dietary Nitrate Intake With Primary Open-Angle Glaucoma: A Prospective Analysis From the Nurses’ Health Study and Health Professionals Follow-up Study. JAMA Ophthalmol. 2016;134:294–303.

19. Kang JH, Willett WC, Rosner BA, et al. Caffeine consumption and the risk of primary open-angle glaucoma: a prospective cohort study. Invest Ophthalmol Vis Sci. 2008;49:1924–1931.

20. Kang JH, Willett WC, Rosner BA, et al. Prospective study of alcohol consumption and the risk of primary open-angle glaucoma. Ophthalmic Epidemiol. 2007;14:141–147.

21. Chiuve SE, Fung TT, Rimm EB, et al. Alternative dietary indices both strongly predict risk of chronic disease. J Nutr. 2012;142:1009–1018.

22. Gao X, Starmer J, Martin ER. A multiple testing correction method for genetic association studies using correlated single nucleotide polymorphisms. Genet Epidemiol. 2008;32:361–369.

23. Benjamini Y, Hochberg Y. Controlling the false discovery rate: a practical and powerful approach to multiple testing. J R Statist Soc B. 1995;57:289–300.

24. Kang JW, Park B, Cho BJ. Comparison of risk factors for initial central scotoma versus initial peripheral scotoma in normal-tension glaucoma. Korean J Ophthalmol. 2015;29:102–108.

25. Kim JM, Kyung H, Shim SH, et al. Location of Initial Visual Field Defects in Glaucoma and Their Modes of Deterioration. Invest Ophthalmol Vis Sci. 2015;56:7956–7962.

26. Park SC, De Moraes CG, Teng CC, et al. Initial parafoveal versus peripheral scotomas in glaucoma: risk factors and visual field characteristics. Ophthalmology. 2011;118:1782–1789.

27. Stegemann C, Pechlaner R, Willeit P, et al. Lipidomics profiling and risk of cardiovascular disease in the prospective population-based Bruneck study. Circulation. 2014;129:1821–1831.

28. Rhee EP, Cheng S, Larson MG, et al. Lipid profiling identifies a triacylglycerol signature of insulin resistance and improves diabetes prediction in humans. J Clin Invest. 2011;121:1402–1411.

29. Umeno A, Tanito M, Kaidzu S, et al. Comprehensive measurements of hydroxylinoleate and hydroxyarachidonate isomers in blood samples from primary open-angle glaucoma patients and controls. Sci Rep. 2019;9:2171.

30. Javadiyan S, Burdon KP, Whiting MJ, et al. Elevation of serum asymmetrical and symmetrical dimethylarginine in patients with advanced glaucoma. Invest Ophthalmol Vis Sci. 2012;53:1923–1927.

31. Gong H, Zhang S, Li Q, et al. Gut microbiota compositional profile and serum metabolic phenotype in patients with primary open-angle glaucoma. Exp Eye Res. 2020;191:107921.

32. Burgess LG, Uppal K, Walker DI, et al. Metabolome-Wide Association Study of Primary Open Angle Glaucoma. Invest Ophthalmol Vis Sci. 2015;56:5020–5028.

33. Leruez S, Marill A, Bresson T, et al. A Metabolomics Profiling of Glaucoma Points to Mitochondrial Dysfunction, Senescence, and Polyamines Deficiency. Invest Ophthalmol Vis Sci. 2018;59:4355–4361.

34. Kouassi Nzoughet J, Guehlouz K, Leruez S, et al. A Data Mining Metabolomics Exploration of Glaucoma. Metabolites. 2020;10.

35. Buisset A, Gohier P, Leruez S, et al. Metabolomic Profiling of Aqueous Humor in Glaucoma Points to Taurine and Spermine Deficiency: Findings from the Eye-D Study. J Proteome Res. 2019;18:1307–1315.

36. Myer C, Perez J, Abdelrahman L, et al. Differentiation of soluble aqueous humor metabolites in primary open angle glaucoma and controls. Exp Eye Res. 2020;194:108024.

37. Pan CW, Ke C, Chen Q, et al. Differential metabolic markers associated with primary open-angle glaucoma and cataract in human aqueous humor. BMC Ophthalmol. 2020;20:183.

38. Cabrerizo J, Urcola JA, Vecino E. Changes in the Lipidomic Profile of Aqueous Humor in Open-Angle Glaucoma. J Glaucoma. 2017;26:349–355.

39. Rossi C, Cicalini I, Cufaro MC, et al. Multi-Omics Approach for Studying Tears in Treatment-Naive Glaucoma Patients. Int J Mol Sci. 2019;20.

40. Boucard CC, Hoogduin JM, van der Grond J, Cornelissen FW. Occipital proton magnetic resonance spectroscopy (1H-MRS) reveals normal metabolite concentrations in retinal visual field defects. PLoS One. 2007;2:e222.

41. Chen YY, Hu HY, Chu D, et al. Patients with Primary Open-Angle Glaucoma May Develop Ischemic Heart Disease More Often than Those without Glaucoma: An 11-Year Population-Based Cohort Study. PLoS One. 2016;11:e0163210.

42. Newman-Casey PA, Talwar N, Nan B, et al. The relationship between components of metabolic syndrome and open-angle glaucoma. Ophthalmology. 2011;118:1318–1326.

43. Xu M, Li S, Zhu J, et al. Plasma lipid levels and risk of primary open angle glaucoma: a genetic study using Mendelian randomization. BMC Ophthalmol. 2020;20:390.

44. Wang S, Bao X. Hyperlipidemia, Blood Lipid Level, and the Risk of Glaucoma: A Meta-Analysis. Invest Ophthalmol Vis Sci. 2019;60:1028–1043.

45. Pertl L, Mossbock G, Wedrich A, et al. Triglycerides and Open Angle Glaucoma - A Meta-analysis with meta-regression. Sci Rep. 2017;7:7829.

46. Rasoulinejad SA, Kasiri A, Montazeri M, et al. The Association Between Primary Open Angle Glaucoma and Clustered Components of Metabolic Syndrome. Open Ophthalmol J. 2015;9:149–155.

47. Anchisi L, Dessi S, Pani A, Mandas A. Cholesterol homeostasis: a key to prevent or slow down neurodegeneration. Front Physiol. 2012;3:486.

48. Huynh K, Lim WLF, Giles C, et al. Concordant peripheral lipidome signatures in two large clinical studies of Alzheimer’s disease. Nat Commun. 2020;11:5698.

49. Llano DA, Devanarayan V, Alzheimer’s Disease Neuroimaging I. Serum Phosphatidylethanolamine and Lysophosphatidylethanolamine Levels Differentiate Alzheimer’s Disease from Controls and Predict Progression from Mild Cognitive Impairment. J Alzheimers Dis. 2021;80:311–319.

50. Wood PL, Locke VA, Herling P, et al. Targeted lipidomics distinguishes patient subgroups in mild cognitive impairment (MCI) and late onset Alzheimer’s disease (LOAD). BBA Clin. 2016;5:25–28.

51. Meikle PJ, Wong G, Barlow CK, et al. Plasma lipid profiling shows similar associations with prediabetes and type 2 diabetes. PLoS One. 2013;8:e74341.

52. Mejia EM, Hatch GM. Mitochondrial phospholipids: role in mitochondrial function. J Bioenerg Biomembr. 2016;48:99–112.

53. Liu P, Zhu W, Chen C, et al. The mechanisms of lysophosphatidylcholine in the development of diseases. Life Sci. 2020;247:117443.

54. Leruez S, Bresson T, Chao de la Barca JM, et al. A Plasma Metabolomic Signature of the Exfoliation Syndrome Involves Amino Acids, Acylcarnitines, and Polyamines. Invest Ophthalmol Vis Sci. 2018;59:1025–1032.

55. Ho LTY, Osterwald A, Ruf I, et al. Role of the autotaxin-lysophosphatidic acid axis in glaucoma, aqueous humor drainage and fibrogenic activity. Biochim Biophys Acta Mol Basis Dis. 2020;1866:165560.

56. Honjo M, Igarashi N, Kurano M, et al. Autotaxin-Lysophosphatidic Acid Pathway in Intraocular Pressure Regulation and Glaucoma Subtypes. Invest Ophthalmol Vis Sci. 2018;59:693–701.

57. Milbeck SM, Bhattacharya SK. Alteration in Lysophospholipids and Converting Enzymes in Glaucomatous Optic Nerves. Invest Ophthalmol Vis Sci. 2020;61:60.

58. Edwards G, Aribindi K, Guerra Y, et al. Phospholipid profiles of control and glaucomatous human aqueous humor. Biochimie. 2014;101:232–247.

59. Aribindi K, Guerra Y, Lee RK, Bhattacharya SK. Comparative phospholipid profiles of control and glaucomatous human trabecular meshwork. Invest Ophthalmol Vis Sci. 2013;54:3037–3044.

60. Kamel K, Farrell M, O’Brien C. Mitochondrial dysfunction in ocular disease: Focus on glaucoma. Mitochondrion. 2017;35:44–53.

61. Liu H, Mercieca K, Prokosch V. Mitochondrial Markers in Aging and Primary Open-Angle Glaucoma. J Glaucoma. 2020;29:295–303.

62. Kong GY, Van Bergen NJ, Trounce IA, Crowston JG. Mitochondrial dysfunction and glaucoma. J Glaucoma. 2009;18:93–100.

63. Chrysostomou V, Rezania F, Trounce IA, Crowston JG. Oxidative stress and mitochondrial dysfunction in glaucoma. Curr Opin Pharmacol. 2013;13:12–15.

64. Izzotti A, Bagnis A, Sacca SC. The role of oxidative stress in glaucoma. Mutat Res. 2006;612:105–114.

65. Benoist d’Azy C, Pereira B, Chiambaretta F, Dutheil F. Oxidative and Anti-Oxidative Stress Markers in Chronic Glaucoma: A Systematic Review and Meta-Analysis. PLoS One. 2016;11:e0166915.

66. Williams PA, Harder JM, Foxworth NE, et al. Vitamin B3 modulates mitochondrial vulnerability and prevents glaucoma in aged mice. Science. 2017;355:756–760.

67. Gonzalez-Granda A, Damms-Machado A, Basrai M, Bischoff SC. Changes in Plasma Acylcarnitine and Lysophosphatidylcholine Levels Following a High-Fructose Diet: A Targeted Metabolomics Study in Healthy Women. Nutrients. 2018;10.

68. Calandrella N, De Seta C, Scarsella G, Risuleo G. Carnitine reduces the lipoperoxidative damage of the membrane and apoptosis after induction of cell stress in experimental glaucoma. Cell Death Dis. 2010;1:e62.

69. Rossi C, Cicalini I, Cufaro MC, et al. Multi-Omics Approach for Studying Tears in Treatment-Naive Glaucoma Patients. Int J Mol Sci. 2019;20:4029.

70. Zarnowski T, Tulidowicz-Bielak M, Kosior-Jarecka E, et al. A ketogenic diet may offer neuroprotection in glaucoma and mitochondrial diseases of the optic nerve. Med Hypothesis Discov Innov Ophthalmol. 2012;1:45–49.

71. Bailey JN, Yaspan BL, Pasquale LR, et al. Hypothesis-independent pathway analysis implicates GABA and acetyl-CoA metabolism in primary open-angle glaucoma and normal-pressure glaucoma. Hum Genet. 2014;133:1319–1330.

72. Hanyuda A, Rosner BA, Wiggs JL, et al. Low-carbohydrate-diet scores and the risk of primary open-angle glaucoma: data from three US cohorts. Eye (Lond). 2020;34:1465–1475.

73. Khawaja AP, Cooke Bailey JN, Kang JH, et al. Assessing the Association of Mitochondrial Genetic Variation With Primary Open-Angle Glaucoma Using Gene-Set Analyses. Invest Ophthalmol Vis Sci. 2016;57:5046–5052.

