## Supplemental material for "Plasma metabolomics of primary open-angle glaucoma in three prospective US cohorts and the UK Biobank"

**Table of Contents**

[**Supplementary Figure S1. Secondary analysis by age (< vs. ≥58.42 years; n=594 vs. n=604).**	3](#_Toc96588039)

[**Supplementary Figure S3. Secondary analysis by BMI (< vs. ≥25 kg/m^2^; n=649 vs. 549)**	5](#_Toc96588041)

[**Supplementary Figure S4. Secondary analysis by time to diagnosis (<11.75 vs. ≥11.75 years; n=598 vs. 600).**	6](#_Toc96588042)

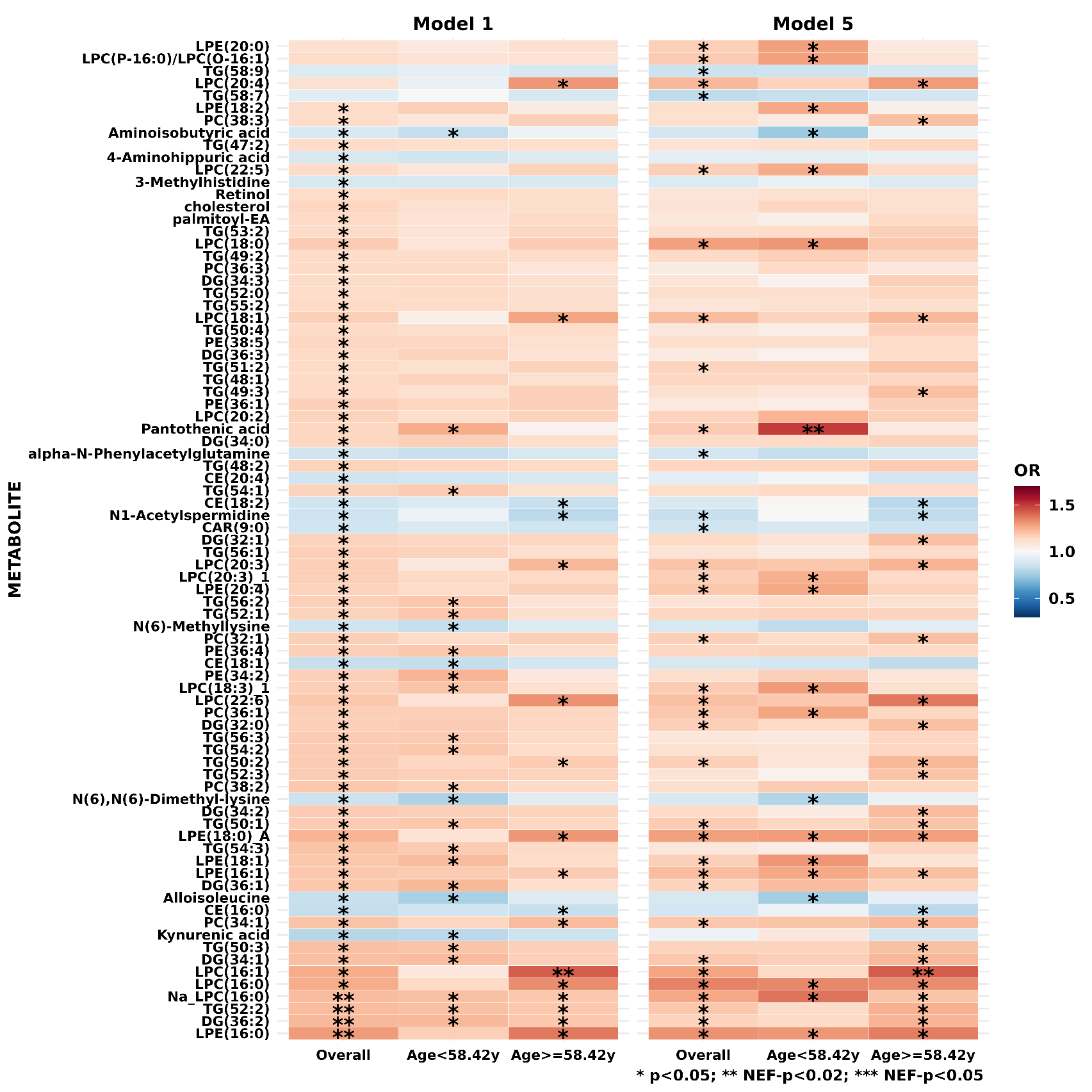

### **Supplementary Figure S1. Secondary analysis by age (< vs. ≥58.42 years; n=594 vs. n=604).**

Metabolites that are nominally significant in either Model 1 or Model 5 are plotted. **Model 1**: basic model, adjusting for matching factors only; **Model 5**: age + smoking status + BMI + physical activity + time of day (as matching imperfect) +month of blood draw (season, as matching imperfect) + family history of POAG + SES + race + age at menopause + nitrate intake + caffeine intake + alcohol intake + alternate healthy eating index + caloric intake + hypertension + high cholesterol + diabetes + oral/inhaled steroid use.

*p<0.05. ** Number of effective tests corrected (NEF)-p<0.2. *** NEF-p<0.05.

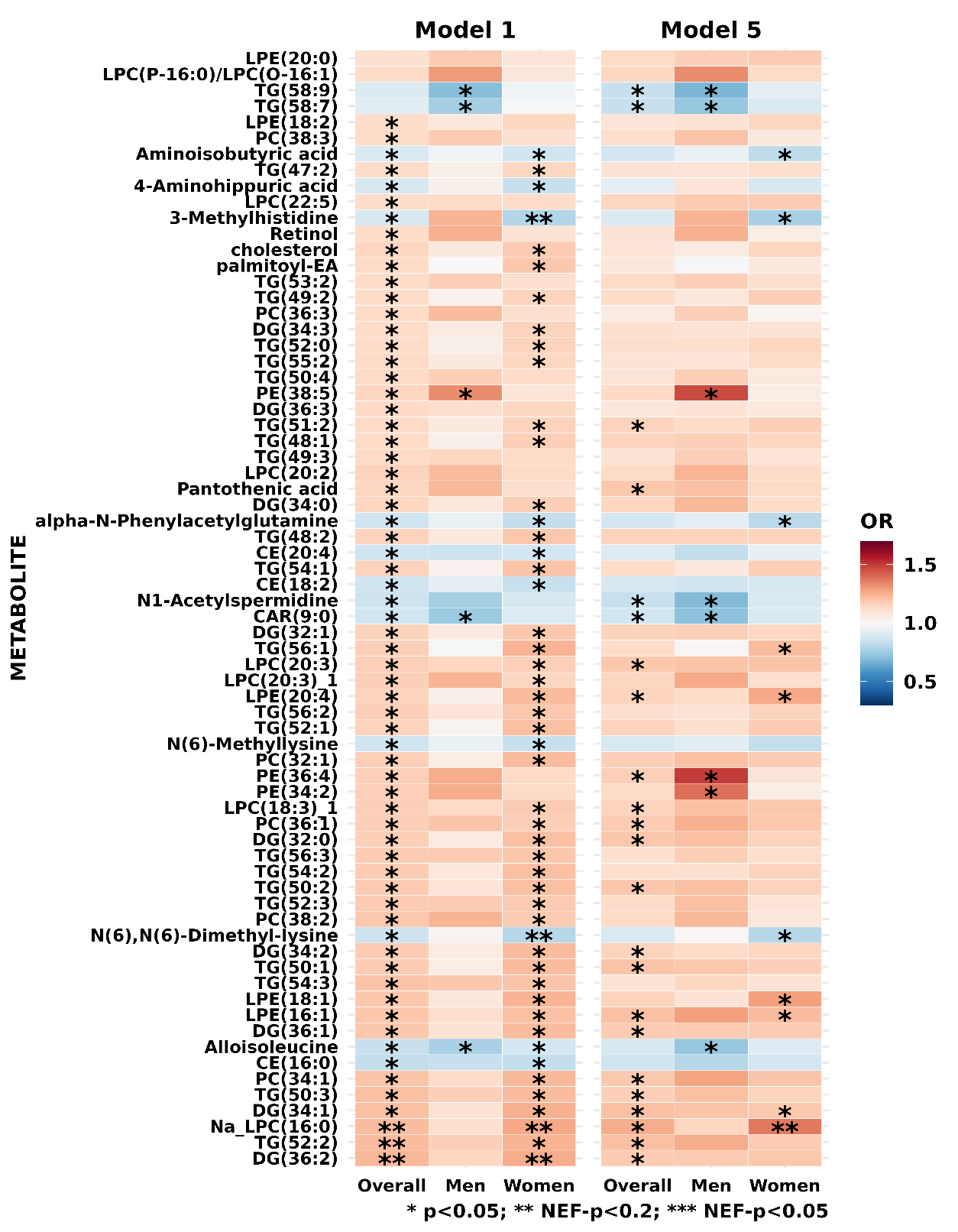

### **Supplementary Figure S2. Secondary analysis by sex (men vs. women; n=308 vs. 890).**

Metabolites that are nominally significant in either Model 1 or Model 5 are plotted. **Model 1**: basic model, adjusting for matching factors only; **Model 5**: age + smoking status + BMI + physical activity + time of day (as matching imperfect) +month of blood draw (season, as matching imperfect) + family history of POAG + SES + race + age at menopause + nitrate intake + caffeine intake + alcohol intake + alternate healthy eating index + caloric intake + hypertension + high cholesterol + diabetes + oral/inhaled steroid use.

*p<0.05. ** Number of effective tests corrected (NEF)-p<0.2. *** NEF-p<0.05.

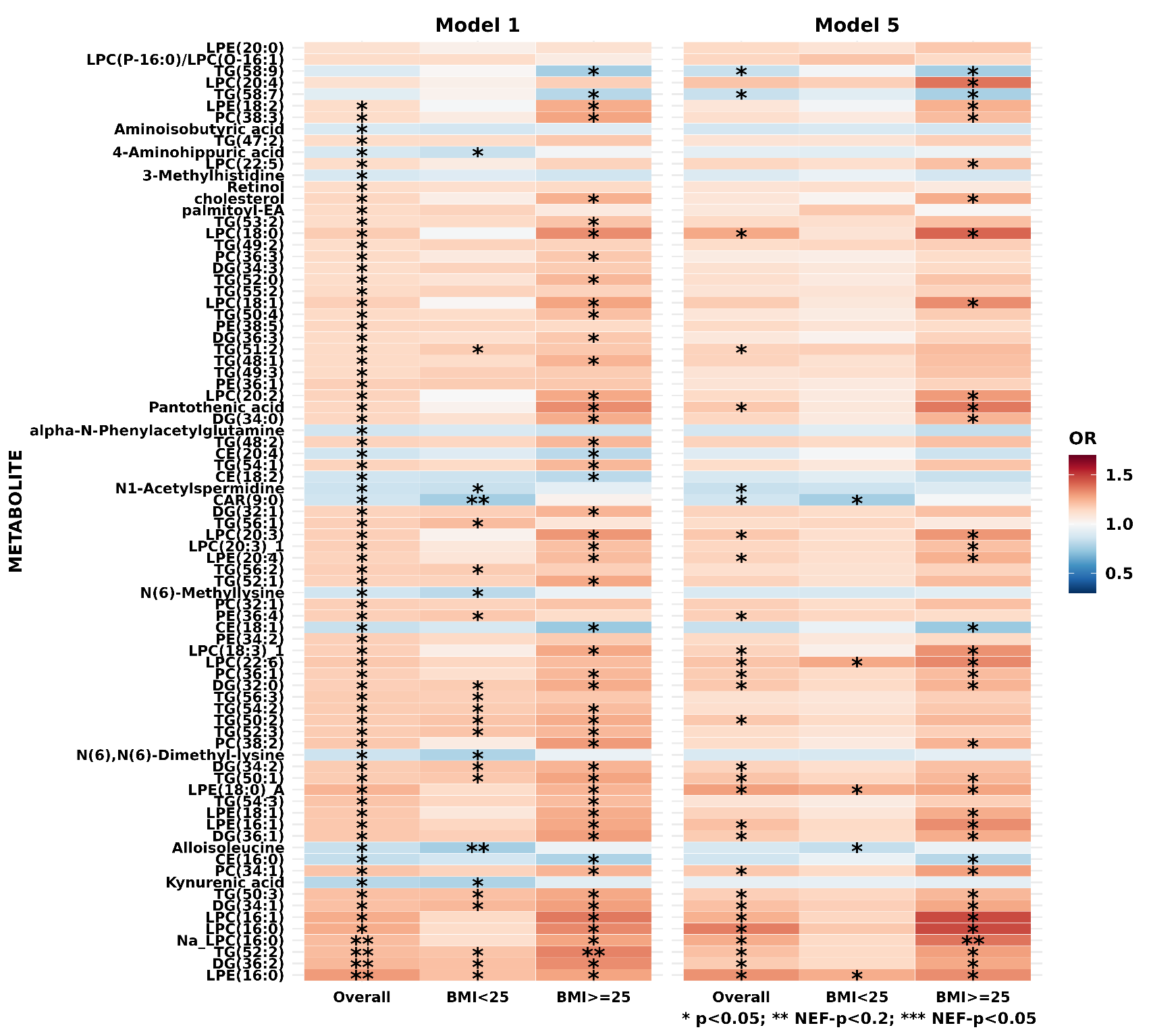

### **Supplementary Figure S3. Secondary analysis by BMI (< vs. ≥25 kg/m^2^; n=649 vs. 549)**

Metabolites that are nominally significant in either Model 1 or Model 5 are plotted. **Model 1**: basic model, adjusting for matching factors only; **Model 5**: age + smoking status + BMI + physical activity + time of day (as matching imperfect) +month of blood draw (season, as matching imperfect) + family history of POAG + SES + race + age at menopause + nitrate intake + caffeine intake + alcohol intake + alternate healthy eating index + caloric intake + hypertension + high cholesterol + diabetes + oral/inhaled steroid use.

*p<0.05. ** Number of effective tests corrected (NEF)-p<0.2. *** NEF-p<0.05.

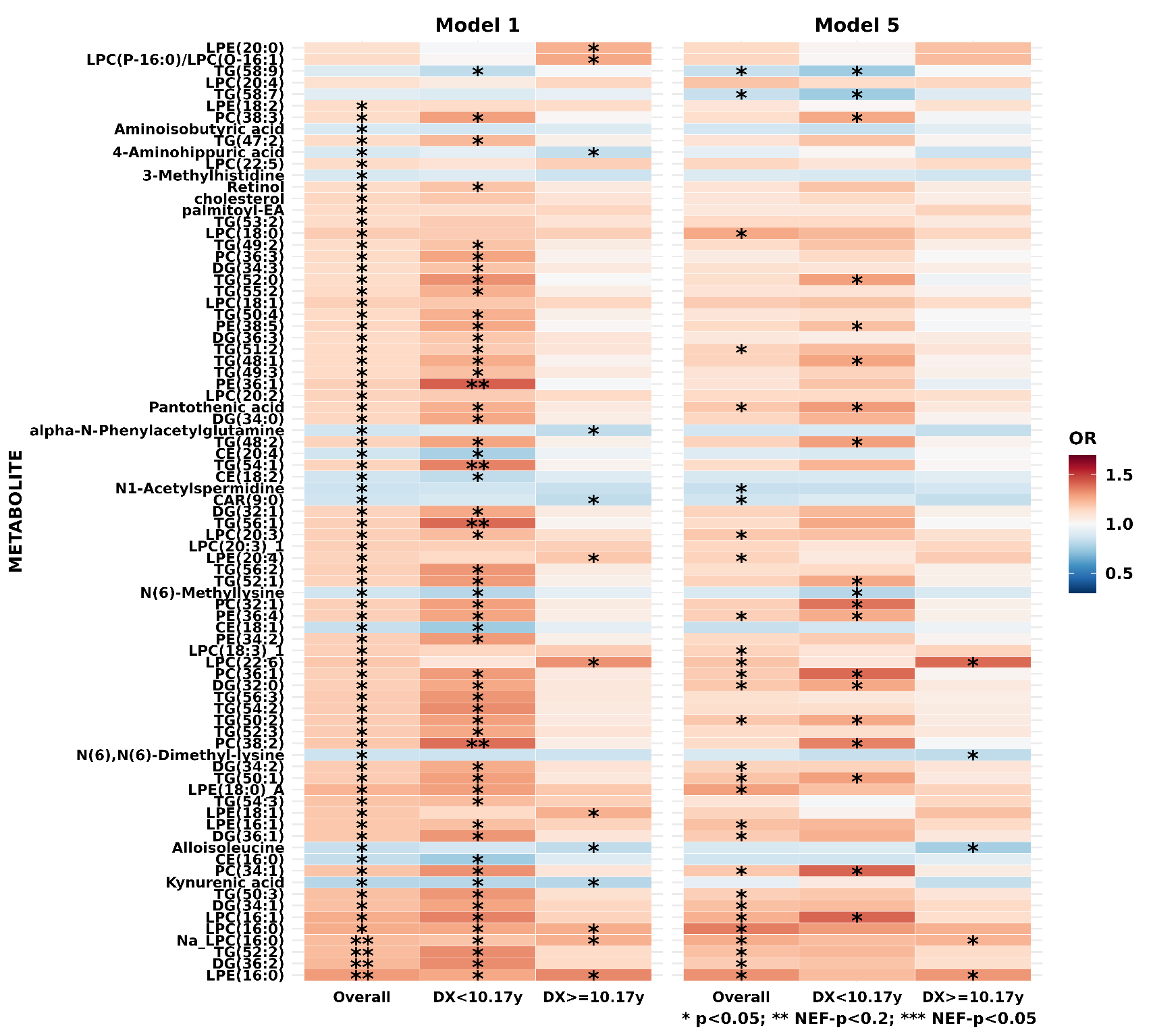

### **Supplementary Figure S4. Secondary analysis by time to diagnosis (<11.75 vs. ≥11.75 years; n=598 vs. 600).**

Metabolites that are nominally significant in either Model 1 or Model 5 are plotted. **Model 1**: basic model, adjusting for matching factors only; **Model 5**: age + smoking status + BMI + physical activity + time of day (as matching imperfect) +month of blood draw (season, as matching imperfect) + family history of POAG + SES + race + age at menopause + nitrate intake + caffeine intake + alcohol intake + alternate healthy eating index + caloric intake + hypertension + high cholesterol + diabetes + oral/inhaled steroid use.

*p<0.05. ** Number of effective tests corrected (NEF)-p<0.2. *** NEF-p<0.05.

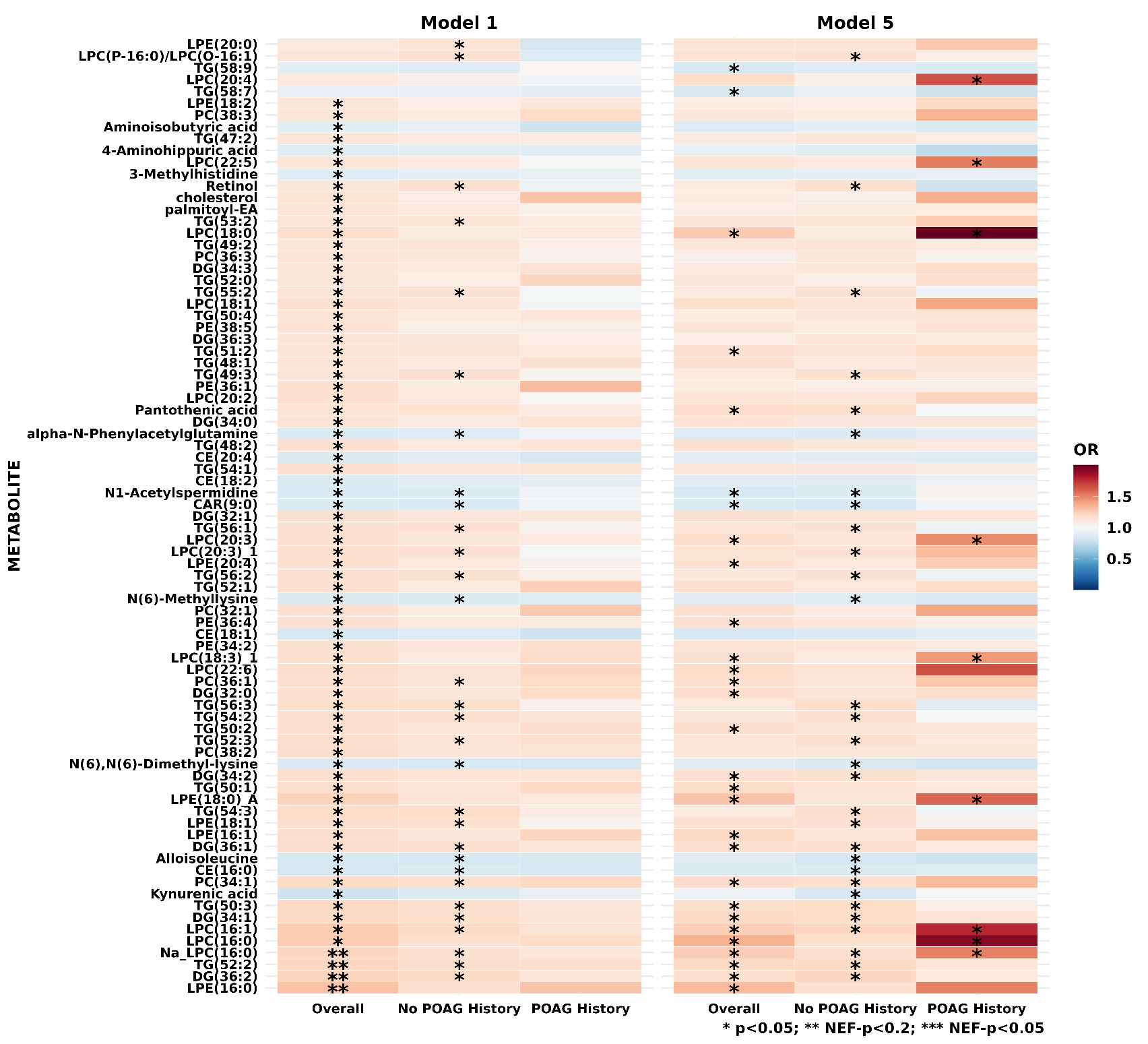

### **Supplementary Figure S5. Secondary analysis by self-reported glaucoma family history (yes vs. no; n=261 vs n=882).**

Metabolites that are nominally significant in either Model 1 or Model 5 are plotted. **Model 1**: basic model, adjusting for matching factors only; **Model 5**: age + smoking status + BMI + physical activity + time of day (as matching imperfect) +month of blood draw (season, as matching imperfect) + family history of POAG + SES + race + age at menopause + nitrate intake + caffeine intake + alcohol intake + alternate healthy eating index + caloric intake + hypertension + high cholesterol + diabetes + oral/inhaled steroid use.

*p<0.05. ** Number of effective tests corrected (NEF)-p<0.2. *** NEF-p<0.05.

### **Supplementary Table S1. Odds ratios (OR) and 95% confidence intervals (CI) of POAG for all metabolites in Model 1 and Model 5 in NHS, NHS2, HPFS (599 POAG cases and 599 controls**

|  |  |  |  | **Model 1** | | | **Model 5** | | |
| --- | --- | --- | --- | --- | --- | --- | --- | --- | --- |
| **HMDB_ID** | **METABOLITE** | **class** | **CV** | **OR (95% CI)** | **p** | **NEF-p** | **OR (95%CI)** | **p** | **NEF-p** |
| HMDB0010382 | LPC(16:0) | Lysophosphatidylcholines | 7.23 | 1.26 (1.09-1.46) | <0.01 | 0.23 | 1.36 (1.12-1.65) | <0.01 | 0.28 |
| NA | Na_LPC(16:0) | Lysophosphatidylcholines | 6.04 | 1.22 (1.08-1.38) | <0.01 | 0.19 | 1.26 (1.08-1.47) | <0.01 | 0.38 |
| HMDB0011503 | LPE(16:0) | Lysophosphatidylethanolamines | 8.25 | 1.30 (1.12-1.50) | <0.01 | 0.07 | 1.32 (1.09-1.60) | <0.01 | 0.63 |
| HMDB0005369* | TG(52:2) | Triglycerides | 6.72 | 1.22 (1.08-1.38) | <0.01 | 0.16 | 1.21 (1.04-1.41) | 0.01 | >0.99 |
| HMDB0007102* | DG(34:1) | Diglycerides | 7.50 | 1.21 (1.07-1.36) | <0.01 | 0.23 | 1.21 (1.04-1.41) | 0.01 | >0.99 |
| HMDB0011504* | LPE(16:1) | Lysophosphatidylethanolamines | 21.38 | 1.19 (1.05-1.34) | <0.01 | 0.64 | 1.21 (1.04-1.40) | 0.01 | >0.99 |
| HMDB0011130 | LPE(18:0)_A | Lysophosphatidylethanolamines | 10.46 | 1.24 (1.06-1.44) | 0.01 | 0.72 | 1.29 (1.05-1.58) | 0.01 | >0.99 |
| HMDB0010383* | LPC(16:1) | Lysophosphatidylcholines | 6.50 | 1.26 (1.09-1.45) | <0.01 | 0.23 | 1.25 (1.04-1.52) | 0.02 | >0.99 |
| HMDB0007972* | PC(34:1) | Phosphatidylcholines | 8.18 | 1.20 (1.06-1.36) | <0.01 | 0.43 | 1.19 (1.03-1.39) | 0.02 | >0.99 |
| HMDB0005360* | TG(50:1) | Triglycerides | 8.90 | 1.18 (1.05-1.33) | 0.01 | 0.78 | 1.20 (1.03-1.40) | 0.02 | >0.99 |
| HMDB0007098* | DG(32:0) | Diglycerides | 11.24 | 1.17 (1.04-1.32) | 0.01 | >0.99 | 1.19 (1.02-1.38) | 0.02 | >0.99 |
| HMDB0000210 | Pantothenic acid | Carboxylic acids and derivatives | 36.16 | 1.15 (1.02-1.29) | 0.02 | >0.99 | 1.19 (1.03-1.37) | 0.02 | >0.99 |
| HMDB0010384 | LPC(18:0) | Lysophosphatidylcholines | 7.69 | 1.18 (1.01-1.37) | 0.04 | >0.99 | 1.27 (1.03-1.55) | 0.02 | >0.99 |
| HMDB0005471* | TG(58:7) | Triglycerides | 13.80 | 0.92 (0.81-1.04) | 0.18 | >0.99 | 0.84 (0.72-0.98) | 0.02 | >0.99 |
| HMDB0007218* | DG(36:2) | Diglycerides | 7.08 | 1.23 (1.09-1.39) | <0.01 | 0.11 | 1.18 (1.02-1.37) | 0.03 | >0.99 |
| HMDB0007216* | DG(36:1) | Diglycerides | 10.70 | 1.19 (1.05-1.34) | <0.01 | 0.61 | 1.18 (1.02-1.37) | 0.03 | >0.99 |
| HMDB0005377* | TG(50:2) | Triglycerides | 9.24 | 1.18 (1.05-1.33) | 0.01 | 0.97 | 1.19 (1.02-1.38) | 0.03 | >0.99 |
| HMDB0008038* | PC(36:1) | Phosphatidylcholines | 8.57 | 1.17 (1.04-1.33) | 0.01 | >0.99 | 1.18 (1.02-1.37) | 0.03 | >0.99 |
| HMDB0010393* | LPC(20:3) | Lysophosphatidylcholines | 7.07 | 1.17 (1.03-1.32) | 0.01 | >0.99 | 1.19 (1.02-1.38) | 0.03 | >0.99 |
| HMDB0001276 | N1-Acetylspermidine | Organic acids and derivatives | 15.12 | 0.85 (0.75-0.97) | 0.02 | >0.99 | 0.84 (0.72-0.98) | 0.03 | >0.99 |
| HMDB0005463* | TG(58:9) | Triglycerides | 8.73 | 0.90 (0.80-1.01) | 0.08 | >0.99 | 0.84 (0.73-0.98) | 0.03 | >0.99 |
| HMDB0005433* | TG(50:3) | Triglycerides | 5.68 | 1.21 (1.07-1.36) | <0.01 | 0.29 | 1.17 (1.01-1.36) | 0.04 | >0.99 |
| HMDB0008937* | PE(36:4) | Phosphatidylethanolamines | 9.87 | 1.17 (1.03-1.32) | 0.01 | >0.99 | 1.17 (1.01-1.35) | 0.04 | >0.99 |
| HMDB0011478* | LPC(18:3)_1 | Lysophosphatidylcholines | 14.71 | 1.17 (1.03-1.31) | 0.01 | >0.99 | 1.16 (1.00-1.35) | 0.04 | >0.99 |
| HMDB0013288 | CAR(9:0) | Carnitines | 24.34 | 0.86 (0.76-0.97) | 0.02 | >0.99 | 0.86 (0.74-0.99) | 0.04 | >0.99 |
| HMDB0005362* | TG(51:2) | Triglycerides | 6.46 | 1.14 (1.01-1.28) | 0.03 | >0.99 | 1.16 (1.00-1.35) | 0.04 | >0.99 |
| HMDB0000212 | N-Acetyl-D-galactosamine | Organic oxygen compounds | 17.94 | 0.89 (0.78-1.00) | 0.05 | >0.99 | 0.85 (0.73-0.99) | 0.04 | >0.99 |
| HMDB0012097 | SM(d18:1/14:0) | Sphingomyelins | 8.27 | 0.92 (0.80-1.06) | 0.25 | >0.99 | 0.82 (0.68-0.99) | 0.04 | >0.99 |
| HMDB0007103* | DG(34:2) | Diglycerides | 8.33 | 1.18 (1.05-1.33) | 0.01 | 0.85 | 1.16 (1.00-1.35) | 0.05 | >0.99 |
| HMDB0000918* | CE(18:1) | Cholesteryl esters | 4.16 | 0.84 (0.73-0.96) | 0.01 | >0.99 | 0.84 (0.70-1.00) | 0.05 | >0.99 |
| HMDB0007099* | DG(32:1) | Diglycerides | 8.72 | 1.16 (1.03-1.31) | 0.01 | >0.99 | 1.16 (1.00-1.35) | 0.05 | >0.99 |
| HMDB0007873* | PC(32:1) | Phosphatidylcholines | 7.95 | 1.17 (1.03-1.32) | 0.01 | >0.99 | 1.17 (1.00-1.36) | 0.05 | >0.99 |
| HMDB0010404 | LPC(22:6) | Lysophosphatidylcholines | 9.01 | 1.19 (1.04-1.36) | 0.01 | >0.99 | 1.20 (1.00-1.43) | 0.05 | >0.99 |
| HMDB0011517 | LPE(20:4) | Lysophosphatidylethanolamines | 7.48 | 1.16 (1.03-1.31) | 0.01 | >0.99 | 1.16 (1.00-1.35) | 0.05 | >0.99 |
| HMDB0010395 | LPC(20:4) | Lysophosphatidylcholines | 8.73 | 1.11 (0.96-1.27) | 0.15 | >0.99 | 1.20 (1.00-1.45) | 0.05 | >0.99 |
| HMDB0000885 | CE(16:0) | Cholesteryl esters | 4.21 | 0.83 (0.74-0.94) | <0.01 | 0.48 | 0.86 (0.74-1.00) | 0.06 | >0.99 |
| HMDB0000557 | Alloisoleucine | NA | 32.83 | 0.84 (0.75-0.95) | <0.01 | 0.50 | 0.88 (0.76-1.00) | 0.06 | >0.99 |
| HMDB0011506* | LPE(18:1) | Lysophosphatidylethanolamines | 6.91 | 1.19 (1.05-1.35) | 0.01 | 0.68 | 1.16 (0.99-1.35) | 0.06 | >0.99 |
| HMDB0005367* | TG(52:1) | Triglycerides | 9.52 | 1.16 (1.03-1.31) | 0.01 | >0.99 | 1.16 (1.00-1.35) | 0.06 | >0.99 |
| HMDB0005376* | TG(48:2) | Triglycerides | 7.71 | 1.16 (1.03-1.30) | 0.02 | >0.99 | 1.16 (1.00-1.35) | 0.06 | >0.99 |
| HMDB0006344 | alpha-N-Phenylacetylglutamine | Organic acids and derivatives | 26.45 | 0.86 (0.77-0.97) | 0.02 | >0.99 | 0.87 (0.76-1.00) | 0.06 | >0.99 |
| HMDB0005359* | TG(48:1) | Triglycerides | 9.87 | 1.14 (1.02-1.29) | 0.03 | >0.99 | 1.16 (0.99-1.35) | 0.06 | >0.99 |
| HMDB0001906 | Aminoisobutyric acid | Organic acids and derivatives | 22.11 | 0.89 (0.78-1.00) | 0.05 | >0.99 | 0.87 (0.75-1.01) | 0.06 | >0.99 |
| HMDB0005478* | TG(60:12) | Triglycerides | 11.48 | 0.90 (0.80-1.01) | 0.08 | >0.99 | 0.87 (0.75-1.00) | 0.06 | >0.99 |
| HMDB0005476* | TG(58:10) | Triglycerides | 5.02 | 0.90 (0.80-1.02) | 0.09 | >0.99 | 0.87 (0.75-1.00) | 0.06 | >0.99 |
| HMDB0000699 | 1-Methyl nicotinamide | Pyridines and derivatives | 30.23 | 1.09 (0.97-1.23) | 0.16 | >0.99 | 1.16 (0.99-1.34) | 0.06 | >0.99 |
| HMDB0007100* | DG(34:0) | Diglycerides | 12.26 | 1.15 (1.02-1.30) | 0.02 | >0.99 | 1.15 (0.99-1.34) | 0.07 | >0.99 |
| HMDB0010403* | LPC(22:5) | Lysophosphatidylcholines | 14.93 | 1.13 (1.00-1.28) | 0.04 | >0.99 | 1.15 (0.99-1.33) | 0.07 | >0.99 |
| HMDB0010407* | LPC(P-16:0)/LPC(O-16:1) | Phosphatidylcholine plasmalogens | 15.05 | 1.13 (0.99-1.28) | 0.07 | >0.99 | 1.15 (0.99-1.34) | 0.07 | >0.99 |
| HMDB0005862 | 2-Methylguanosine | NA | 24.93 | 0.90 (0.80-1.02) | 0.09 | >0.99 | 0.87 (0.76-1.01) | 0.07 | >0.99 |
| HMDB0000532 | N-Acetylglycine | NA | 64.97 | 0.91 (0.80-1.03) | 0.15 | >0.99 | 0.87 (0.75-1.01) | 0.07 | >0.99 |
| HMDB0010394* | LPC(20:3)_1 | Lysophosphatidylcholines | 22.80 | 1.17 (1.03-1.33) | 0.01 | >0.99 | 1.15 (0.99-1.34) | 0.08 | >0.99 |
| HMDB0009069* | PE(38:5) | Phosphatidylethanolamines | 28.10 | 1.15 (1.01-1.29) | 0.03 | >0.99 | 1.14 (0.99-1.31) | 0.08 | >0.99 |
| HMDB0005357* | TG(50:0) | Triglycerides | 13.19 | 1.12 (1.00-1.26) | 0.05 | >0.99 | 1.15 (0.99-1.33) | 0.08 | >0.99 |
| HMDB0005392* | TG(56:8) | Triglycerides | 3.86 | 0.92 (0.81-1.03) | 0.14 | >0.99 | 0.88 (0.76-1.01) | 0.08 | >0.99 |
| HMDB0000897 | 7-Methylguanine | Organoheterocyclic compounds | 15.31 | 0.92 (0.81-1.04) | 0.17 | >0.99 | 0.87 (0.75-1.02) | 0.08 | >0.99 |
| HMDB0000462 | Allantoin | Azoles | 38.11 | 1.05 (0.92-1.19) | 0.47 | >0.99 | 1.15 (0.99-1.34) | 0.08 | >0.99 |
| HMDB0008928* | PE(34:2) | Phosphatidylethanolamines | 13.12 | 1.17 (1.04-1.32) | 0.01 | >0.99 | 1.14 (0.98-1.31) | 0.09 | >0.99 |
| HMDB0000610* | CE(18:2) | Cholesteryl esters | 4.11 | 0.86 (0.77-0.97) | 0.02 | >0.99 | 0.88 (0.76-1.02) | 0.09 | >0.99 |
| HMDB0010392 | LPC(20:2) | Lysophosphatidylcholines | 17.25 | 1.16 (1.02-1.31) | 0.02 | >0.99 | 1.14 (0.98-1.33) | 0.09 | >0.99 |
| HMDB0002815* | LPC(18:1) | Lysophosphatidylcholines | 6.65 | 1.17 (1.01-1.36) | 0.03 | >0.99 | 1.18 (0.97-1.43) | 0.09 | >0.99 |
| HMDB0011706* | TG(49:2) | Triglycerides | 7.35 | 1.13 (1.01-1.27) | 0.04 | >0.99 | 1.13 (0.98-1.31) | 0.09 | >0.99 |
| HMDB0042196* | TG(53:2) | Triglycerides | 7.53 | 1.13 (1.01-1.28) | 0.04 | >0.99 | 1.14 (0.98-1.32) | 0.09 | >0.99 |
| HMDB0002172 | N1,N12-Diacetylspermine | Organic acids and derivatives | 65.65 | 0.95 (0.84-1.06) | 0.35 | >0.99 | 0.88 (0.76-1.02) | 0.09 | >0.99 |
| HMDB0002038 | N(6)-Methyllysine | NA | 32.03 | 0.86 (0.77-0.97) | 0.01 | >0.99 | 0.89 (0.77-1.02) | 0.10 | >0.99 |
| HMDB0013272 | N-Lauroylglycine | NA | 45.26 | 0.88 (0.76-1.02) | 0.10 | >0.99 | 0.86 (0.72-1.03) | 0.10 | >0.99 |
| HMDB0011481 | LPE(20:0) | Lysophosphatidylethanolamines | 3.83 | 1.11 (0.98-1.26) | 0.11 | >0.99 | 1.14 (0.97-1.33) | 0.10 | >0.99 |
| HMDB0000991 | 2-Aminooctanoic acid | Organic acids and derivatives | 16.80 | 0.91 (0.81-1.03) | 0.13 | >0.99 | 0.89 (0.77-1.02) | 0.10 | >0.99 |
| HMDB0013287 | N(6),N(6)-Dimethyl-lysine | Organic acids and derivatives | 20.55 | 0.85 (0.76-0.96) | 0.01 | 0.87 | 0.89 (0.77-1.03) | 0.11 | >0.99 |
| HMDB0013130 | CAR(DC5:0) | Carnitines | 17.16 | 0.89 (0.79-1.00) | 0.06 | >0.99 | 0.89 (0.77-1.03) | 0.11 | >0.99 |
| HMDB0005356* | TG(48:0) | Triglycerides | 10.80 | 1.12 (0.99-1.25) | 0.07 | >0.99 | 1.13 (0.97-1.32) | 0.11 | >0.99 |
| HMDB0006733 | CE(22:6) | Cholesteryl esters | 7.90 | 0.90 (0.80-1.01) | 0.07 | >0.99 | 0.89 (0.76-1.03) | 0.11 | >0.99 |
| HMDB0042104* | TG(51:1) | Triglycerides | 8.66 | 1.10 (0.98-1.24) | 0.10 | >0.99 | 1.12 (0.97-1.30) | 0.11 | >0.99 |
| HMDB0008952* | PE(P-34:1)/PE(O-34:2) | Phosphatidylethanolamine plasmalogens | 13.03 | 1.07 (0.95-1.21) | 0.25 | >0.99 | 1.13 (0.97-1.31) | 0.11 | >0.99 |
| HMDB0005384* | TG(52:3) | Triglycerides | 6.11 | 1.18 (1.05-1.33) | 0.01 | 0.95 | 1.13 (0.97-1.31) | 0.12 | >0.99 |
| HMDB0005404* | TG(56:2) | Triglycerides | 17.54 | 1.17 (1.03-1.32) | 0.01 | >0.99 | 1.12 (0.97-1.30) | 0.12 | >0.99 |
| HMDB0005395* | TG(54:1) | Triglycerides | 17.25 | 1.16 (1.03-1.30) | 0.02 | >0.99 | 1.13 (0.97-1.31) | 0.12 | >0.99 |
| NA | PC(12:0/12:0) | Phosphatidylcholines | 4.42 | 1.17 (0.98-1.39) | 0.07 | >0.99 | 1.17 (0.96-1.43) | 0.12 | >0.99 |
| HMDB0011508* | LPE(18:3) | Lysophosphatidylethanolamines | 15.17 | 1.11 (0.99-1.26) | 0.08 | >0.99 | 1.13 (0.97-1.31) | 0.12 | >0.99 |
| HMDB0008270* | PC(38:2) | Phosphatidylcholines | 8.89 | 1.19 (1.05-1.36) | 0.01 | 0.90 | 1.13 (0.96-1.31) | 0.13 | >0.99 |
| HMDB0005403* | TG(54:2) | Triglycerides | 10.17 | 1.18 (1.05-1.34) | 0.01 | 0.97 | 1.12 (0.97-1.31) | 0.13 | >0.99 |
| HMDB0005396* | TG(56:1) | Triglycerides | 22.19 | 1.17 (1.03-1.34) | 0.01 | >0.99 | 1.13 (0.96-1.32) | 0.13 | >0.99 |
| HMDB0005365* | TG(52:0) | Triglycerides | 20.10 | 1.13 (1.01-1.28) | 0.04 | >0.99 | 1.12 (0.97-1.31) | 0.13 | >0.99 |
| HMDB0000982 | 5-Methylcytidine | NA | 14.99 | 0.89 (0.79-1.00) | 0.05 | >0.99 | 0.89 (0.77-1.03) | 0.13 | >0.99 |
| HMDB0010412* | TG(46:1) | Triglycerides | 11.78 | 1.12 (1.00-1.26) | 0.06 | >0.99 | 1.12 (0.97-1.31) | 0.13 | >0.99 |
| HMDB0010411* | TG(46:0) | Triglycerides | 12.95 | 1.12 (0.99-1.26) | 0.07 | >0.99 | 1.13 (0.97-1.32) | 0.13 | >0.99 |
| HMDB0001932 | Metoprolol | NA | 94.20 | 0.86 (0.70-1.07) | 0.18 | >0.99 | 0.81 (0.62-1.07) | 0.14 | >0.99 |
| HMDB0000479 | 3-Methylhistidine | Organic acids and derivatives | 16.46 | 0.88 (0.78-1.00) | 0.04 | >0.99 | 0.90 (0.78-1.04) | 0.15 | >0.99 |
| HMDB0007132* | DG(34:3) | Diglycerides | 7.32 | 1.13 (1.01-1.28) | 0.04 | >0.99 | 1.11 (0.96-1.29) | 0.15 | >0.99 |
| HMDB0008047* | PC(38:3) | Phosphatidylcholines | 8.53 | 1.13 (1.00-1.27) | 0.05 | >0.99 | 1.12 (0.96-1.29) | 0.15 | >0.99 |
| HMDB0042100* | TG(47:1) | Triglycerides | 11.55 | 1.12 (1.00-1.26) | 0.05 | >0.99 | 1.11 (0.96-1.29) | 0.15 | >0.99 |
| HMDB0010379 | LPC(14:0) | Lysophosphatidylcholines | 7.07 | 1.14 (1.00-1.31) | 0.06 | >0.99 | 1.15 (0.95-1.39) | 0.15 | >0.99 |
| NA | PS(P-36:2)/PS(O-36:3) | Phosphatidylserine plasmalogens | 19.74 | 1.12 (1.00-1.26) | 0.06 | >0.99 | 1.11 (0.96-1.28) | 0.15 | >0.99 |
| HMDB0000033 | Carnosine | NA | 55.24 | 1.09 (0.96-1.25) | 0.19 | >0.99 | 1.13 (0.96-1.33) | 0.15 | >0.99 |
| HMDB0000182 | Lysine | Carboxylic acids and derivatives | 14.11 | 1.06 (0.94-1.20) | 0.35 | >0.99 | 1.11 (0.96-1.29) | 0.16 | >0.99 |
| HMDB0004827 | Proline betaine | Organic acids and derivatives | 48.03 | 0.96 (0.86-1.08) | 0.53 | >0.99 | 0.90 (0.78-1.04) | 0.16 | >0.99 |
| HMDB0000305 | Retinol | NA | 21.25 | 1.13 (1.01-1.28) | 0.04 | >0.99 | 1.10 (0.96-1.27) | 0.17 | >0.99 |
| HMDB0000641 | Glutamine | Organic acids and derivatives | 12.82 | 0.90 (0.79-1.03) | 0.11 | >0.99 | 0.89 (0.76-1.05) | 0.17 | >0.99 |
| HMDB0005410* | TG(56:3) | Triglycerides | 13.65 | 1.18 (1.05-1.34) | 0.01 | >0.99 | 1.11 (0.95-1.29) | 0.18 | >0.99 |
| HMDB0042103* | TG(49:3) | Triglycerides | 7.69 | 1.14 (1.02-1.28) | 0.03 | >0.99 | 1.10 (0.96-1.28) | 0.18 | >0.99 |
| HMDB0000875 | Trigonelline | Alkaloids and derivatives | 14.46 | 0.92 (0.82-1.04) | 0.20 | >0.99 | 0.90 (0.77-1.05) | 0.18 | >0.99 |
| HMDB0042076* | TG(47:2) | Triglycerides | 13.12 | 1.13 (1.00-1.26) | 0.04 | >0.99 | 1.10 (0.95-1.27) | 0.19 | >0.99 |
| HMDB0005385* | TG(54:5) | Triglycerides | 34.00 | 1.09 (0.97-1.23) | 0.13 | >0.99 | 1.10 (0.95-1.26) | 0.19 | >0.99 |
| HMDB0042226* | TG(55:2) | Triglycerides | 18.13 | 1.14 (1.01-1.28) | 0.04 | >0.99 | 1.10 (0.95-1.27) | 0.20 | >0.99 |
| HMDB0009003* | PE(38:4) | Phosphatidylethanolamines | 8.97 | 1.12 (0.99-1.26) | 0.06 | >0.99 | 1.10 (0.95-1.26) | 0.20 | >0.99 |
| HMDB0004193 | N1-Methyl-2-pyridone-5-carboxamide | Organoheterocyclic compounds | 38.82 | 1.05 (0.93-1.18) | 0.41 | >0.99 | 1.10 (0.95-1.27) | 0.20 | >0.99 |
| HMDB0000824 | CAR(3:0) | Carnitines | 20.87 | 1.00 (0.89-1.13) | 0.95 | >0.99 | 1.10 (0.95-1.28) | 0.20 | >0.99 |
| HMDB0006726 | CE(20:4) | Cholesteryl esters | 4.86 | 0.86 (0.76-0.97) | 0.02 | >0.99 | 0.91 (0.79-1.05) | 0.21 | >0.99 |
| HMDB0010391 | LPC(20:1) | Lysophosphatidylcholines | 7.43 | 1.10 (0.97-1.24) | 0.13 | >0.99 | 1.10 (0.95-1.28) | 0.21 | >0.99 |
| HMDB0005448* | TG(56:9) | Triglycerides | 7.71 | 0.94 (0.84-1.06) | 0.29 | >0.99 | 0.91 (0.80-1.05) | 0.21 | >0.99 |
| HMDB0010531* | TG(58:11) | Triglycerides | 10.73 | 0.92 (0.82-1.03) | 0.15 | >0.99 | 0.92 (0.80-1.05) | 0.22 | >0.99 |
| HMDB0002869 | campesterol | NA | 23.94 | 1.13 (0.99-1.29) | 0.08 | >0.99 | 1.11 (0.94-1.30) | 0.23 | >0.99 |
| NA | PC(36:4-OH) | Phosphatidylcholines | 19.70 | 1.12 (0.99-1.26) | 0.08 | >0.99 | 1.09 (0.94-1.27) | 0.23 | >0.99 |
| HMDB0010419* | TG(46:2) | Triglycerides | 15.93 | 1.11 (0.98-1.25) | 0.09 | >0.99 | 1.09 (0.94-1.27) | 0.23 | >0.99 |
| HMDB0005462* | TG(56:7) | Triglycerides | 6.15 | 0.97 (0.86-1.09) | 0.57 | >0.99 | 0.92 (0.79-1.06) | 0.23 | >0.99 |
| NA | Ectoine | NA | 59.42 | 1.03 (0.91-1.17) | 0.66 | >0.99 | 1.10 (0.94-1.30) | 0.23 | >0.99 |
| HMDB0005405* | TG(54:3) | Triglycerides | 6.38 | 1.20 (1.05-1.36) | 0.01 | 0.71 | 1.10 (0.94-1.28) | 0.24 | >0.99 |
| HMDB0005432* | TG(48:3) | Triglycerides | 11.35 | 1.12 (1.00-1.27) | 0.05 | >0.99 | 1.09 (0.94-1.27) | 0.24 | >0.99 |
| HMDB0042099* | TG(45:1) | Triglycerides | 19.09 | 1.12 (1.00-1.26) | 0.06 | >0.99 | 1.09 (0.94-1.26) | 0.24 | >0.99 |
| HMDB0005435* | TG(50:4) | Triglycerides | 8.65 | 1.14 (1.01-1.29) | 0.03 | >0.99 | 1.09 (0.94-1.26) | 0.25 | >0.99 |
| HMDB0007870* | PC(30:1) | Phosphatidylcholines | 9.11 | 1.09 (0.97-1.23) | 0.14 | >0.99 | 1.09 (0.94-1.27) | 0.25 | >0.99 |
| HMDB0013238 | CAR(7:0) | Carnitines | 13.95 | 0.92 (0.82-1.04) | 0.21 | >0.99 | 0.92 (0.79-1.06) | 0.25 | >0.99 |
| HMDB0013713 | N-Acetyl-D-tryptophan | Organic acids and derivatives | 27.31 | 1.06 (0.94-1.19) | 0.35 | >0.99 | 1.09 (0.94-1.25) | 0.25 | >0.99 |
| HMDB0000895 | Acetylcholine | Organonitrogen compounds | 70.31 | 1.00 (0.88-1.15) | 0.95 | >0.99 | 0.90 (0.76-1.08) | 0.25 | >0.99 |
| HMDB0000067 | cholesterol | NA | 11.47 | 1.15 (1.01-1.30) | 0.04 | >0.99 | 1.09 (0.94-1.27) | 0.26 | >0.99 |
| HMDB0011507* | LPE(18:2) | Lysophosphatidylethanolamines | 8.44 | 1.13 (1.00-1.28) | 0.05 | >0.99 | 1.09 (0.94-1.28) | 0.26 | >0.99 |
| HMDB0000562 | Creatinine | Carboxylic acids and derivatives | 11.14 | 0.91 (0.80-1.03) | 0.13 | >0.99 | 0.92 (0.79-1.06) | 0.26 | >0.99 |
| HMDB0005458* | TG(58:6) | Triglycerides | 7.35 | 0.95 (0.84-1.07) | 0.37 | >0.99 | 0.92 (0.80-1.06) | 0.26 | >0.99 |
| HMDB0000062 | Carnitine | Carnitines | 11.93 | 1.00 (0.88-1.13) | 0.95 | >0.99 | 1.09 (0.94-1.28) | 0.26 | >0.99 |
| HMDB0007219* | DG(36:3) | Diglycerides | 6.50 | 1.14 (1.01-1.29) | 0.03 | >0.99 | 1.08 (0.94-1.25) | 0.27 | >0.99 |
| NA | C-Glycosyltryptophan | NA | 23.43 | 0.89 (0.79-1.02) | 0.09 | >0.99 | 0.92 (0.79-1.07) | 0.27 | >0.99 |
| HMDB0042063* | TG(44:0) | Triglycerides | 19.48 | 1.10 (0.98-1.24) | 0.12 | >0.99 | 1.09 (0.94-1.27) | 0.27 | >0.99 |
| HMDB0011520 | LPE(22:0) | Lysophosphatidylethanolamines | 9.19 | 1.07 (0.94-1.21) | 0.30 | >0.99 | 1.09 (0.94-1.27) | 0.27 | >0.99 |
| HMDB0011243* | PC(P-36:1)/PC(O-36:2) | Phosphatidylcholine plasmalogens | 8.24 | 0.99 (0.88-1.11) | 0.82 | >0.99 | 0.92 (0.80-1.07) | 0.27 | >0.99 |
| HMDB0008993* | PE(36:1) | Phosphatidylethanolamines | 12.37 | 1.17 (1.02-1.35) | 0.02 | >0.99 | 1.10 (0.93-1.31) | 0.28 | >0.99 |
| HMDB0002100 | palmitoyl-EA | NA | 5.66 | 1.14 (1.01-1.29) | 0.04 | >0.99 | 1.08 (0.93-1.25) | 0.29 | >0.99 |
| HMDB0006736* | CE(20:3) | Cholesteryl esters | 8.57 | 0.89 (0.79-1.00) | 0.05 | >0.99 | 0.93 (0.80-1.07) | 0.29 | >0.99 |
| HMDB0042301* | TG(44:1) | Triglycerides | 20.25 | 1.10 (0.98-1.24) | 0.10 | >0.99 | 1.08 (0.93-1.26) | 0.29 | >0.99 |
| HMDB0007871* | PC(32:0) | Phosphatidylcholines | 8.72 | 1.08 (0.96-1.22) | 0.20 | >0.99 | 1.09 (0.93-1.26) | 0.29 | >0.99 |
| HMDB0000658* | CE(16:1) | Cholesteryl esters | 5.38 | 1.07 (0.95-1.21) | 0.23 | >0.99 | 1.08 (0.93-1.25) | 0.29 | >0.99 |
| HMDB0000766 | N-Acetylalanine | Organic acids and derivatives | 12.67 | 1.03 (0.91-1.17) | 0.63 | >0.99 | 1.08 (0.93-1.26) | 0.30 | >0.99 |
| HMDB0054096 | TG(54:10) | Triglycerides | 9.63 | 1.06 (0.94-1.20) | 0.33 | >0.99 | 1.08 (0.93-1.24) | 0.31 | >0.99 |
| HMDB0001867 | 4-Aminohippuric acid | NA | 30.82 | 0.88 (0.78-1.00) | 0.04 | >0.99 | 0.93 (0.80-1.08) | 0.32 | >0.99 |
| HMDB0010390 | LPC(20:0) | Lysophosphatidylcholines | 8.96 | 1.06 (0.94-1.20) | 0.31 | >0.99 | 1.08 (0.93-1.27) | 0.32 | >0.99 |
| HMDB0000168 | Asparagine | Organic acids and derivatives | 10.69 | 0.95 (0.84-1.07) | 0.38 | >0.99 | 0.93 (0.80-1.08) | 0.32 | >0.99 |
| HMDB0013127 | CAR(4:0(OH)) | Carnitines | 27.11 | 0.96 (0.85-1.09) | 0.52 | >0.99 | 0.93 (0.80-1.08) | 0.32 | >0.99 |
| HMDB0031106* | TG(51:0) | Triglycerides | 21.58 | 1.09 (0.97-1.23) | 0.13 | >0.99 | 1.08 (0.93-1.25) | 0.34 | >0.99 |
| HMDB0000904 | Citrulline | Organic acids and derivatives | 15.37 | 0.92 (0.81-1.04) | 0.16 | >0.99 | 0.93 (0.80-1.08) | 0.34 | >0.99 |
| HMDB0000925 | Trimethylamine N-oxide | Organonitrogen compounds | 33.87 | 0.94 (0.83-1.06) | 0.31 | >0.99 | 0.93 (0.81-1.08) | 0.34 | >0.99 |
| NA | N-Methyl-proline | NA | 48.67 | 0.97 (0.86-1.09) | 0.62 | >0.99 | 0.93 (0.80-1.08) | 0.34 | >0.99 |
| HMDB0007011* | DG(30:0) | Diglycerides | 20.21 | 1.14 (0.97-1.34) | 0.11 | >0.99 | 1.09 (0.90-1.32) | 0.35 | >0.99 |
| HMDB0007199* | DG(38:5) | Diglycerides | 6.70 | 1.09 (0.97-1.23) | 0.13 | >0.99 | 1.07 (0.93-1.23) | 0.35 | >0.99 |
| HMDB0002250 | CAR(12:0) | Carnitines | 16.84 | 0.91 (0.80-1.03) | 0.14 | >0.99 | 0.93 (0.80-1.08) | 0.35 | >0.99 |
| HMDB0000679 | Homocitrulline | Organic acids and derivatives | 37.70 | 0.93 (0.83-1.05) | 0.27 | >0.99 | 0.93 (0.80-1.08) | 0.35 | >0.99 |
| HMDB0003157 | Guanidinosuccinic acid | NA | 33.86 | 1.07 (0.94-1.22) | 0.28 | >0.99 | 1.08 (0.92-1.27) | 0.35 | >0.99 |
| HMDB0002820 | Methylimidazoleacetic acid | Organoheterocyclic compounds | 30.56 | 0.92 (0.82-1.04) | 0.19 | >0.99 | 0.94 (0.81-1.08) | 0.36 | >0.99 |
| HMDB0006455 | CAR(20:4) | Carnitines | 63.22 | 1.04 (0.92-1.18) | 0.56 | >0.99 | 1.07 (0.92-1.24) | 0.36 | >0.99 |
| HMDB0008994* | PE(36:2) | Phosphatidylethanolamines | 9.42 | 1.13 (1.00-1.27) | 0.05 | >0.99 | 1.07 (0.92-1.23) | 0.37 | >0.99 |
| HMDB0001539 | Dimethylarginine | Organic acids and derivatives | 16.79 | 0.88 (0.76-1.01) | 0.07 | >0.99 | 0.92 (0.77-1.10) | 0.38 | >0.99 |
| HMDB0042751* | TG(46:3) | Triglycerides | 18.32 | 1.11 (0.99-1.25) | 0.08 | >0.99 | 1.07 (0.92-1.24) | 0.38 | >0.99 |
| HMDB0013331* | CAR(14:2) | Carnitines | 15.93 | 0.92 (0.81-1.04) | 0.19 | >0.99 | 0.94 (0.81-1.09) | 0.38 | >0.99 |
| HMDB0003681 | 4-Acetamidobutanoic acid | Organic acids and derivatives | 11.52 | 1.05 (0.93-1.18) | 0.43 | >0.99 | 1.07 (0.92-1.23) | 0.38 | >0.99 |
| HMDB0009082* | PE(P-36:1)/PE(O-36:2) | Phosphatidylethanolamine plasmalogens | 9.56 | 1.05 (0.93-1.18) | 0.45 | >0.99 | 1.07 (0.92-1.23) | 0.38 | >0.99 |
| HMDB0010397 | LPC(20:5) | Lysophosphatidylcholines | 8.13 | 1.02 (0.91-1.16) | 0.70 | >0.99 | 1.07 (0.92-1.25) | 0.39 | >0.99 |
| HMDB0008942* | PE(38:2) | Phosphatidylethanolamines | 8.75 | 0.98 (0.87-1.10) | 0.76 | >0.99 | 0.94 (0.81-1.09) | 0.39 | >0.99 |
| HMDB0002005 | Methionine sulfoxide | NA | 24.87 | 0.95 (0.84-1.08) | 0.43 | >0.99 | 0.94 (0.81-1.09) | 0.40 | >0.99 |
| HMDB0029377 | Piperine | Alkaloids and derivatives | 40.82 | 0.97 (0.86-1.10) | 0.64 | >0.99 | 0.94 (0.81-1.09) | 0.40 | >0.99 |
| HMDB0003334 | Symmetric dimethyl-arginine | Organic acids and derivatives | 18.90 | 0.92 (0.81-1.05) | 0.23 | >0.99 | 0.94 (0.80-1.09) | 0.41 | >0.99 |
| HMDB0013733 | Trimethylbenzene | Benzene and substituted derivatives | 23.36 | 0.94 (0.82-1.08) | 0.37 | >0.99 | 0.93 (0.79-1.10) | 0.41 | >0.99 |
| HMDB0005456* | TG(56:6) | Triglycerides | 5.24 | 0.96 (0.85-1.08) | 0.46 | >0.99 | 0.94 (0.82-1.08) | 0.41 | >0.99 |
| NA | phenylalanine-d8 [iSTD] | NA | 6.52 | 0.95 (0.83-1.09) | 0.50 | >0.99 | 0.93 (0.80-1.10) | 0.41 | >0.99 |
| HMDB0007869* | PC(30:0) | Phosphatidylcholines | 9.33 | 1.06 (0.94-1.20) | 0.32 | >0.99 | 1.06 (0.92-1.23) | 0.42 | >0.99 |
| HMDB0010471* | TG(50:5) | Triglycerides | 12.15 | 1.09 (0.97-1.23) | 0.14 | >0.99 | 1.06 (0.92-1.23) | 0.43 | >0.99 |
| HMDB0032055 | N-Acetylhistidine | NA | 17.02 | 0.93 (0.82-1.05) | 0.22 | >0.99 | 0.94 (0.81-1.09) | 0.43 | >0.99 |
| HMDB0011387* | PE(P-38:5)/PE(O-38:6) | Phosphatidylethanolamine plasmalogens | 7.96 | 1.00 (0.89-1.13) | 0.94 | >0.99 | 1.06 (0.92-1.22) | 0.43 | >0.99 |
| HMDB0000898 | 1-Methylhistamine | NA | 33.41 | 0.92 (0.81-1.05) | 0.21 | >0.99 | 0.94 (0.81-1.10) | 0.44 | >0.99 |
| HMDB0011701* | TG(51:3) | Triglycerides | 7.54 | 1.10 (0.98-1.24) | 0.10 | >0.99 | 1.06 (0.91-1.23) | 0.45 | >0.99 |
| HMDB0010498* | TG(54:9) | Triglycerides | 34.70 | 0.95 (0.85-1.07) | 0.44 | >0.99 | 0.95 (0.82-1.09) | 0.46 | >0.99 |
| HMDB0008138* | PC(36:4)_B | Phosphatidylcholines | 6.66 | 1.04 (0.92-1.17) | 0.54 | >0.99 | 1.06 (0.91-1.22) | 0.47 | >0.99 |
| HMDB0008105* | PC(36:3) | Phosphatidylcholines | 8.06 | 1.14 (1.01-1.30) | 0.04 | >0.99 | 1.06 (0.91-1.23) | 0.48 | >0.99 |
| HMDB0001563 | 1-Methylguanosine | NA | 16.81 | 0.92 (0.82-1.04) | 0.21 | >0.99 | 0.95 (0.82-1.10) | 0.48 | >0.99 |
| HMDB0000802 | Pterin | NA | 40.07 | 1.05 (0.93-1.18) | 0.46 | >0.99 | 1.05 (0.91-1.22) | 0.48 | >0.99 |
| HMDB0043170* | TG(45:2) | Triglycerides | 22.88 | 1.10 (0.98-1.24) | 0.10 | >0.99 | 1.05 (0.91-1.22) | 0.49 | >0.99 |
| HMDB0000043 | Betaine | Carboxylic acids and derivatives | 10.86 | 0.95 (0.84-1.08) | 0.43 | >0.99 | 0.94 (0.80-1.11) | 0.49 | >0.99 |
| HMDB0010375* | CE(22:5) | Cholesteryl esters | 19.95 | 0.90 (0.80-1.02) | 0.11 | >0.99 | 0.95 (0.82-1.10) | 0.50 | >0.99 |
| HMDB0072780* | TG(42:0) | Triglycerides | 25.93 | 1.07 (0.95-1.20) | 0.25 | >0.99 | 1.05 (0.91-1.23) | 0.50 | >0.99 |
| HMDB0000054 | Bilirubin | NA | 66.73 | 1.04 (0.92-1.18) | 0.51 | >0.99 | 1.05 (0.90-1.23) | 0.50 | >0.99 |
| HMDB0002271 | Imidazolepropionic acid | Organoheterocyclic compounds | 27.84 | 1.03 (0.91-1.15) | 0.66 | >0.99 | 1.05 (0.92-1.19) | 0.50 | >0.99 |
| HMDB0001847 | Caffeine | Imidazopyrimidines | 21.10 | 0.99 (0.88-1.12) | 0.93 | >0.99 | 0.95 (0.80-1.12) | 0.51 | >0.99 |
| HMDB0011526 | LPE(22:6) | Lysophosphatidylethanolamines | 10.81 | 1.12 (0.99-1.26) | 0.08 | >0.99 | 1.05 (0.91-1.21) | 0.52 | >0.99 |
| HMDB0000159 | Phenylalanine | Organic acids and derivatives | 15.30 | 0.95 (0.84-1.07) | 0.37 | >0.99 | 0.95 (0.83-1.10) | 0.52 | >0.99 |
| HMDB0000177 | Histidine | Carboxylic acids and derivatives | 16.06 | 1.04 (0.91-1.18) | 0.59 | >0.99 | 1.05 (0.90-1.22) | 0.52 | >0.99 |
| HMDB0042466* | TG(55:3) | Triglycerides | 7.93 | 0.99 (0.88-1.11) | 0.82 | >0.99 | 0.95 (0.82-1.11) | 0.52 | >0.99 |
| HMDB0011410* | PE(P-36:4)/PE(O-36:5) | Phosphatidylethanolamine plasmalogens | 8.30 | 0.99 (0.89-1.12) | 0.93 | >0.99 | 1.05 (0.91-1.21) | 0.52 | >0.99 |
| HMDB0042811* | TG(48:4) | Triglycerides | 15.21 | 1.08 (0.96-1.22) | 0.19 | >0.99 | 1.05 (0.90-1.22) | 0.53 | >0.99 |
| HMDB0009102* | PE(38:6) | Phosphatidylethanolamines | 8.49 | 1.10 (0.97-1.24) | 0.13 | >0.99 | 1.05 (0.90-1.21) | 0.54 | >0.99 |
| HMDB0039059 | 3-Dehydroxycarnitine | NA | 14.86 | 0.91 (0.80-1.03) | 0.14 | >0.99 | 0.95 (0.81-1.11) | 0.54 | >0.99 |
| HMDB0042279* | TG(44:2) | Triglycerides | 21.05 | 1.08 (0.96-1.21) | 0.23 | >0.99 | 1.05 (0.90-1.22) | 0.54 | >0.99 |
| HMDB0000715 | Kynurenic acid | NA | 20.98 | 0.80 (0.69-0.93) | <0.01 | 0.39 | 0.94 (0.78-1.14) | 0.55 | >0.99 |
| NA | PE(P-42:10)/PE(O-42:11) | Phosphatidylethanolamine plasmalogens | 17.65 | 0.99 (0.88-1.12) | 0.92 | >0.99 | 0.96 (0.82-1.11) | 0.55 | >0.99 |
| HMDB0006731 | CE(20:5) | Cholesteryl esters | 6.05 | 0.90 (0.80-1.01) | 0.07 | >0.99 | 0.96 (0.83-1.11) | 0.56 | >0.99 |
| HMDB0000026 | Ureidopropionic acid | Organic acids and derivatives | 17.80 | 0.91 (0.81-1.03) | 0.15 | >0.99 | 0.96 (0.83-1.11) | 0.56 | >0.99 |
| HMDB0042548* | TG(46:4) | Triglycerides | 21.42 | 1.09 (0.97-1.23) | 0.16 | >0.99 | 1.05 (0.90-1.21) | 0.56 | >0.99 |
| HMDB0000161 | Alanine | Carboxylic acids and derivatives | 9.62 | 1.08 (0.96-1.22) | 0.18 | >0.99 | 1.04 (0.91-1.20) | 0.56 | >0.99 |
| HMDB0009016* | PE(P-36:0)/PE(O-36:1) | Phosphatidylethanolamine plasmalogens | 18.25 | 0.98 (0.87-1.11) | 0.77 | >0.99 | 0.96 (0.82-1.11) | 0.56 | >0.99 |
| HMDB0004400 | 5-Acetylamino-6-amino-3-methyluracil | Organic nitrogen compounds | 19.38 | 0.96 (0.86-1.08) | 0.51 | >0.99 | 0.95 (0.81-1.12) | 0.57 | >0.99 |
| HMDB0006725 | CE(14:0) | Cholesteryl esters | 7.78 | 0.98 (0.86-1.13) | 0.82 | >0.99 | 0.95 (0.78-1.14) | 0.57 | >0.99 |
| HMDB0000883 | Valine | Carboxylic acids and derivatives | 13.30 | 1.00 (0.89-1.13) | >0.99 | >0.99 | 1.05 (0.90-1.22) | 0.57 | >0.99 |
| HMDB0042098* | TG(43:1) | Triglycerides | 41.60 | 1.12 (0.98-1.28) | 0.11 | >0.99 | 1.05 (0.89-1.23) | 0.58 | >0.99 |
| HMDB0005380* | TG(52:5) | Triglycerides | 5.96 | 1.10 (0.98-1.23) | 0.12 | >0.99 | 1.04 (0.90-1.21) | 0.59 | >0.99 |
| HMDB0010386* | LPC(18:2) | Lysophosphatidylcholines | 7.99 | 1.04 (0.92-1.18) | 0.53 | >0.99 | 1.05 (0.89-1.23) | 0.59 | >0.99 |
| HMDB0000128 | Guanidoacetic acid | Carboxylic acids and derivatives | 25.69 | 0.97 (0.85-1.09) | 0.58 | >0.99 | 0.96 (0.82-1.12) | 0.59 | >0.99 |
| HMDB0000812 | N-Acetylaspartic acid | Organic acids and derivatives | 10.63 | 0.98 (0.86-1.11) | 0.75 | >0.99 | 0.96 (0.83-1.11) | 0.59 | >0.99 |
| HMDB0000289 | Uric acid | Imidazopyrimidines | 21.87 | 1.00 (0.88-1.13) | 0.99 | >0.99 | 1.04 (0.90-1.21) | 0.59 | >0.99 |
| HMDB0008006* | PC(34:3) | Phosphatidylcholines | 7.67 | 1.13 (1.00-1.28) | 0.05 | >0.99 | 1.04 (0.90-1.20) | 0.60 | >0.99 |
| HMDB0000112 | gamma-Aminobutyric acid | Organic acids and derivatives | 11.70 | 0.95 (0.83-1.09) | 0.46 | >0.99 | 0.95 (0.80-1.14) | 0.61 | >0.99 |
| HMDB0000631* | Glycodeoxycholic acid/Glycochenodeoxycholic acid | NA | 20.59 | 0.98 (0.87-1.10) | 0.72 | >0.99 | 0.96 (0.84-1.11) | 0.61 | >0.99 |
| HMDB0000929 | Tryptophan | Organoheterocyclic compounds | 15.04 | 0.99 (0.88-1.11) | 0.84 | >0.99 | 1.04 (0.90-1.20) | 0.61 | >0.99 |
| HMDB0002802 | Cortisone | Steroids and steroid derivatives | 20.65 | 1.00 (0.88-1.14) | 0.97 | >0.99 | 1.04 (0.89-1.22) | 0.61 | >0.99 |
| HMDB0004824 | N2,N2-Dimethylguanosine | Nucleosides, nucleotides, and analogues | 14.42 | 0.94 (0.83-1.06) | 0.33 | >0.99 | 0.96 (0.83-1.11) | 0.62 | >0.99 |
| HMDB0005370* | TG(54:4) | Triglycerides | 6.22 | 1.06 (0.93-1.19) | 0.39 | >0.99 | 0.96 (0.82-1.12) | 0.62 | >0.99 |
| HMDB0000884 | Ribothymidine | Nucleosides, nucleotides, and analogues | 22.80 | 0.98 (0.87-1.11) | 0.76 | >0.99 | 1.04 (0.90-1.20) | 0.62 | >0.99 |
| HMDB0011343* | PE(P-34:2)/PE(O-34:3) | Phosphatidylethanolamine plasmalogens | 11.24 | 1.01 (0.90-1.13) | 0.89 | >0.99 | 1.04 (0.90-1.19) | 0.62 | >0.99 |
| HMDB0008039* | PC(36:2) | Phosphatidylcholines | 7.30 | 1.09 (0.96-1.23) | 0.18 | >0.99 | 1.04 (0.89-1.21) | 0.63 | >0.99 |
| HMDB0042789* | TG(48:5) | Triglycerides | 17.29 | 1.07 (0.95-1.20) | 0.27 | >0.99 | 1.04 (0.89-1.20) | 0.63 | >0.99 |
| HMDB0009012* | PE(40:6) | Phosphatidylethanolamines | 14.37 | 1.09 (0.97-1.24) | 0.16 | >0.99 | 1.03 (0.90-1.20) | 0.64 | >0.99 |
| NA | TG(45:3) | Triglycerides | 25.60 | 1.08 (0.95-1.24) | 0.24 | >0.99 | 1.04 (0.88-1.23) | 0.64 | >0.99 |
| HMDB0008048* | PC(38:4) | Phosphatidylcholines | 6.92 | 1.04 (0.92-1.17) | 0.54 | >0.99 | 1.03 (0.90-1.19) | 0.64 | >0.99 |
| HMDB0000248 | Thyroxine | Carboxylic acids and derivatives | 16.96 | 0.97 (0.85-1.10) | 0.64 | >0.99 | 0.96 (0.82-1.13) | 0.64 | >0.99 |
| HMDB0000767 | Pseudouridine | Nucleosides, nucleotides, and analogues | 31.52 | 1.00 (0.89-1.13) | 0.99 | >0.99 | 1.03 (0.90-1.19) | 0.64 | >0.99 |
| HMDB0005363* | TG(52:4) | Triglycerides | 5.31 | 1.09 (0.97-1.23) | 0.14 | >0.99 | 1.04 (0.89-1.20) | 0.65 | >0.99 |
| HMDB0000791 | CAR(8:0) | Carnitines | 21.00 | 0.94 (0.83-1.07) | 0.35 | >0.99 | 0.97 (0.83-1.12) | 0.65 | >0.99 |
| HMDB0011420* | PE(P-38:6)/PE(O-38:7) | Phosphatidylethanolamine plasmalogens | 8.70 | 0.97 (0.86-1.09) | 0.57 | >0.99 | 0.97 (0.84-1.12) | 0.65 | >0.99 |
| HMDB0010513* | TG(56:10) | Triglycerides | 16.18 | 0.96 (0.84-1.10) | 0.59 | >0.99 | 0.96 (0.81-1.14) | 0.65 | >0.99 |
| HMDB0000235 | Thiamine | NA | 14.62 | 1.02 (0.90-1.14) | 0.79 | >0.99 | 1.03 (0.89-1.19) | 0.65 | >0.99 |
| HMDB0006347 | CAR(26:0) | Carnitines | 17.23 | 0.99 (0.88-1.12) | 0.89 | >0.99 | 0.97 (0.84-1.11) | 0.65 | >0.99 |
| HMDB0011386* | PE(P-38:4)/PE(O-38:5) | Phosphatidylethanolamine plasmalogens | 8.22 | 1.01 (0.89-1.13) | 0.93 | >0.99 | 1.03 (0.89-1.20) | 0.65 | >0.99 |
| HMDB0010368 | CE(18:0) | Cholesteryl esters | 5.68 | 0.93 (0.83-1.05) | 0.26 | >0.99 | 0.97 (0.83-1.12) | 0.66 | >0.99 |
| HMDB0012107* | SM(d18:1/24:1) | Sphingomyelins | 8.65 | 1.05 (0.93-1.18) | 0.43 | >0.99 | 1.03 (0.89-1.19) | 0.66 | >0.99 |
| HMDB0011103 | 1,7-Dimethyluric acid | Organoheterocyclic compounds | 24.20 | 0.98 (0.87-1.10) | 0.74 | >0.99 | 0.97 (0.83-1.13) | 0.66 | >0.99 |
| HMDB0001008 | Biliverdin | Organoheterocyclic compounds | 33.96 | 0.98 (0.87-1.11) | 0.72 | >0.99 | 0.97 (0.83-1.13) | 0.67 | >0.99 |
| HMDB0008057* | PC(40:6) | Phosphatidylcholines | 7.80 | 1.10 (0.97-1.24) | 0.13 | >0.99 | 1.03 (0.89-1.19) | 0.68 | >0.99 |
| HMDB0008511* | PC(40:10) | Phosphatidylcholines | 8.19 | 1.05 (0.94-1.18) | 0.39 | >0.99 | 0.97 (0.84-1.12) | 0.68 | >0.99 |
| HMDB0010518* | TG(54:8) | Triglycerides | 11.78 | 0.99 (0.88-1.12) | 0.91 | >0.99 | 0.97 (0.84-1.12) | 0.68 | >0.99 |
| HMDB0011511 | LPC(20:0)_1 | Lysophosphatidylcholines | 11.63 | 1.04 (0.92-1.18) | 0.53 | >0.99 | 1.03 (0.89-1.19) | 0.69 | >0.99 |
| HMDB0007991* | PC(38:6) | Phosphatidylcholines | 6.69 | 1.03 (0.92-1.16) | 0.57 | >0.99 | 0.97 (0.84-1.12) | 0.69 | >0.99 |
| HMDB0000687 | Leucine | Carboxylic acids and derivatives | 11.92 | 1.02 (0.91-1.15) | 0.75 | >0.99 | 1.03 (0.89-1.19) | 0.69 | >0.99 |
| HMDB0005406* | TG(56:5) | Triglycerides | 5.60 | 0.95 (0.85-1.07) | 0.42 | >0.99 | 0.97 (0.85-1.12) | 0.70 | >0.99 |
| HMDB0000651 | CAR(10:0) | Carnitines | 16.24 | 0.95 (0.84-1.08) | 0.45 | >0.99 | 0.97 (0.84-1.13) | 0.70 | >0.99 |
| HMDB0011394* | PE(P-40:6)/PE(O-40:7) | Phosphatidylethanolamine plasmalogens | 8.02 | 0.99 (0.88-1.11) | 0.81 | >0.99 | 0.97 (0.85-1.12) | 0.71 | >0.99 |
| HMDB0012102 | SM(d18:1/20:0) | Sphingomyelins | 8.42 | 1.01 (0.89-1.14) | 0.93 | >0.99 | 0.97 (0.84-1.13) | 0.71 | >0.99 |
| HMDB0009060* | PE(36:3) | Phosphatidylethanolamines | 10.34 | 1.12 (0.99-1.26) | 0.08 | >0.99 | 1.03 (0.88-1.19) | 0.72 | >0.99 |
| HMDB0011252* | PC(P-38:3)/PC(O-38:4) | Phosphatidylcholine plasmalogens | 8.41 | 1.06 (0.94-1.20) | 0.35 | >0.99 | 1.03 (0.89-1.19) | 0.72 | >0.99 |
| HMDB0000630 | Cytosine | Diazines | 26.81 | 0.95 (0.84-1.08) | 0.46 | >0.99 | 0.97 (0.83-1.14) | 0.72 | >0.99 |
| HMDB0005066 | CAR(14:0) | Carnitines | 18.47 | 0.97 (0.86-1.09) | 0.58 | >0.99 | 0.97 (0.85-1.12) | 0.72 | >0.99 |
| HMDB0000070 | Pipecolic acid | NA | 16.52 | 0.97 (0.86-1.10) | 0.65 | >0.99 | 1.03 (0.89-1.18) | 0.72 | >0.99 |
| HMDB0061384 | Acisoga | NA | 14.44 | 0.94 (0.83-1.06) | 0.32 | >0.99 | 0.97 (0.84-1.13) | 0.73 | >0.99 |
| HMDB0010497* | TG(50:6) | Triglycerides | 15.05 | 1.05 (0.93-1.19) | 0.41 | >0.99 | 1.03 (0.89-1.19) | 0.73 | >0.99 |
| HMDB0008731* | PC(40:9) | Phosphatidylcholines | 4.78 | 1.03 (0.92-1.15) | 0.64 | >0.99 | 0.98 (0.84-1.13) | 0.73 | >0.99 |
| HMDB0000201 | CAR(2:0) | Carnitines | 13.45 | 1.01 (0.90-1.14) | 0.85 | >0.99 | 1.03 (0.89-1.19) | 0.73 | >0.99 |
| HMDB0005436* | TG(52:6) | Triglycerides | 9.29 | 1.06 (0.94-1.19) | 0.35 | >0.99 | 1.02 (0.89-1.18) | 0.74 | >0.99 |
| HMDB0008923* | PE(32:0) | Phosphatidylethanolamines | 18.52 | 1.07 (0.93-1.22) | 0.37 | >0.99 | 1.03 (0.86-1.23) | 0.74 | >0.99 |
| HMDB0011220* | PC(P-36:4)/PC(O-36:5) | Phosphatidylcholine plasmalogens | 7.61 | 0.96 (0.86-1.08) | 0.53 | >0.99 | 1.02 (0.89-1.19) | 0.74 | >0.99 |
| HMDB0001348 | SM(d18:1/18:0) | Sphingomyelins | 9.42 | 1.04 (0.92-1.17) | 0.57 | >0.99 | 1.02 (0.89-1.18) | 0.74 | >0.99 |
| HMDB0005391* | TG(54:6) | Triglycerides | 6.07 | 1.01 (0.90-1.13) | 0.89 | >0.99 | 1.02 (0.89-1.18) | 0.74 | >0.99 |
| HMDB0001390 | trans-3-Hydroxycotinine | Organoheterocyclic compounds | 20.97 | 1.01 (0.89-1.15) | 0.91 | >0.99 | 1.03 (0.88-1.20) | 0.74 | >0.99 |
| HMDB0011512* | LPE(20:1) | Lysophosphatidylethanolamines | 11.25 | 1.03 (0.91-1.17) | 0.62 | >0.99 | 1.03 (0.88-1.19) | 0.75 | >0.99 |
| NA | PS(P-36:1)/PS(O-36:2) | Phosphatidylserine plasmalogens | 18.71 | 1.02 (0.91-1.16) | 0.70 | >0.99 | 1.02 (0.88-1.19) | 0.75 | >0.99 |
| HMDB0003357 | N-Acetylornithine | Organic acids and derivatives | 19.82 | 0.98 (0.87-1.10) | 0.74 | >0.99 | 1.02 (0.89-1.18) | 0.75 | >0.99 |
| HMDB0072022 | TG(41:0) | Triglycerides | 37.80 | 0.99 (0.77-1.28) | 0.96 | >0.99 | 1.05 (0.77-1.45) | 0.75 | >0.99 |
| HMDB0007874* | PC(32:2) | Phosphatidylcholines | 8.59 | 1.04 (0.92-1.18) | 0.50 | >0.99 | 0.98 (0.85-1.13) | 0.76 | >0.99 |
| HMDB0004953* | Cer(d18:1/24:1) | Ceramides | 8.65 | 1.06 (0.94-1.20) | 0.32 | >0.99 | 0.98 (0.84-1.13) | 0.77 | >0.99 |
| HMDB0011208* | PC(P-34:0)/PC(O-34:1)_A | Phosphatidylcholine plasmalogens | 8.65 | 1.03 (0.92-1.16) | 0.63 | >0.99 | 0.98 (0.85-1.13) | 0.77 | >0.99 |
| HMDB0011441* | PE(P-36:2)/PE(O-36:3) | Phosphatidylethanolamine plasmalogens | 10.14 | 1.03 (0.92-1.15) | 0.63 | >0.99 | 1.02 (0.89-1.18) | 0.77 | >0.99 |
| HMDB0007248* | DG(36:4) | Diglycerides | 6.85 | 1.02 (0.91-1.15) | 0.71 | >0.99 | 0.98 (0.85-1.13) | 0.77 | >0.99 |
| HMDB0000162 | Proline | Carboxylic acids and derivatives | 10.37 | 1.03 (0.91-1.16) | 0.64 | >0.99 | 1.02 (0.88-1.18) | 0.78 | >0.99 |
| HMDB0011621 | Cinnamoylglycine | Organic acids and derivatives | 29.34 | 0.98 (0.87-1.10) | 0.68 | >0.99 | 0.98 (0.85-1.13) | 0.78 | >0.99 |
| HMDB0000138 | Glycocholic acid | Steroids and steroid derivatives | 21.03 | 0.98 (0.87-1.10) | 0.75 | >0.99 | 0.98 (0.85-1.13) | 0.78 | >0.99 |
| HMDB0000725 | Hydroxyproline | Carboxylic acids and derivatives | 15.11 | 0.94 (0.84-1.05) | 0.28 | >0.99 | 0.98 (0.86-1.12) | 0.79 | >0.99 |
| HMDB0001046 | Cotinine | Organoheterocyclic compounds | 22.18 | 0.95 (0.83-1.10) | 0.51 | >0.99 | 0.98 (0.81-1.17) | 0.79 | >0.99 |
| HMDB0000172 | Isoleucine | Organic acids and derivatives | 11.84 | 1.02 (0.90-1.15) | 0.73 | >0.99 | 1.02 (0.87-1.19) | 0.79 | >0.99 |
| HMDB0011384* | PE(P-38:2)/PE(O-38:3) | Phosphatidylethanolamine plasmalogens | 10.80 | 0.98 (0.87-1.11) | 0.78 | >0.99 | 0.98 (0.85-1.13) | 0.79 | >0.99 |
| HMDB0007973* | PC(34:2) | Phosphatidylcholines | 6.26 | 1.06 (0.93-1.22) | 0.37 | >0.99 | 0.98 (0.81-1.18) | 0.80 | >0.99 |
| HMDB0000064 | Creatine | Organic acids and derivatives | 15.22 | 0.98 (0.87-1.10) | 0.73 | >0.99 | 0.98 (0.86-1.13) | 0.80 | >0.99 |
| HMDB0011229* | PC(P-38:6)/PC(O-38:7) | Phosphatidylcholine plasmalogens | 8.89 | 0.98 (0.87-1.10) | 0.74 | >0.99 | 0.98 (0.85-1.13) | 0.80 | >0.99 |
| HMDB0000714 | Hippuric acid | Benzene and substituted derivatives | 18.93 | 0.99 (0.87-1.11) | 0.82 | >0.99 | 0.98 (0.85-1.14) | 0.81 | >0.99 |
| HMDB0002013 | CAR(4:0) | Carnitines | 21.74 | 0.91 (0.81-1.02) | 0.11 | >0.99 | 0.98 (0.85-1.14) | 0.82 | >0.99 |
| HMDB0240212 | DMGV | NA | 16.23 | 0.98 (0.87-1.11) | 0.75 | >0.99 | 1.02 (0.87-1.19) | 0.83 | >0.99 |
| HMDB0000063 | Cortisol | Steroids and steroid derivatives | 19.27 | 0.99 (0.88-1.13) | 0.93 | >0.99 | 1.02 (0.88-1.18) | 0.83 | >0.99 |
| HMDB0001072 | coenzyme Q10 | NA | 15.24 | 1.04 (0.92-1.17) | 0.54 | >0.99 | 0.99 (0.85-1.14) | 0.84 | >0.99 |
| HMDB0004949 | Cer(d18:1/16:0) | Ceramides | 10.17 | 1.05 (0.93-1.19) | 0.44 | >0.99 | 0.99 (0.85-1.14) | 0.85 | >0.99 |
| HMDB0042062* | TG(43:0) | Triglycerides | 35.58 | 1.05 (0.91-1.21) | 0.48 | >0.99 | 0.98 (0.82-1.17) | 0.85 | >0.99 |
| HMDB0001886 | 3-Methylxanthine | Organoheterocyclic compounds | 19.17 | 1.01 (0.90-1.13) | 0.86 | >0.99 | 0.99 (0.85-1.14) | 0.85 | >0.99 |
| HMDB0010169 | SM(d18:1/16:0) | Sphingomyelins | 8.75 | 1.03 (0.91-1.17) | 0.61 | >0.99 | 1.01 (0.87-1.18) | 0.87 | >0.99 |
| HMDB0001859 | Acetaminophen | Phenols | 22.29 | 1.02 (0.90-1.16) | 0.72 | >0.99 | 1.01 (0.87-1.18) | 0.87 | >0.99 |
| HMDB0002014* | CAR(14:1) | Carnitines | 16.35 | 0.98 (0.87-1.11) | 0.75 | >0.99 | 0.99 (0.85-1.14) | 0.87 | >0.99 |
| HMDB0001325 | N-6-Trimethyllysine | Organic acids and derivatives | 14.09 | 0.90 (0.80-1.02) | 0.10 | >0.99 | 0.99 (0.86-1.14) | 0.88 | >0.99 |
| HMDB0005398* | TG(56:4) | Triglycerides | 7.48 | 1.07 (0.95-1.20) | 0.29 | >0.99 | 1.01 (0.87-1.17) | 0.88 | >0.99 |
| HMDB0011214* | PC(P-34:4)/PC(O-34:5) | Phosphatidylcholine plasmalogens | 14.62 | 0.95 (0.85-1.07) | 0.42 | >0.99 | 0.99 (0.86-1.14) | 0.88 | >0.99 |
| HMDB0012101* | SM(d18:1/18:1) | Sphingomyelins | 8.65 | 1.03 (0.90-1.19) | 0.65 | >0.99 | 1.01 (0.84-1.23) | 0.88 | >0.99 |
| HMDB0000670 | Homoarginine | Organic acids and derivatives | 24.31 | 1.02 (0.90-1.15) | 0.76 | >0.99 | 1.01 (0.87-1.17) | 0.88 | >0.99 |
| HMDB0004030 | 21-Deoxycortisol | NA | 29.63 | 0.99 (0.87-1.12) | 0.85 | >0.99 | 1.01 (0.87-1.17) | 0.88 | >0.99 |
| HMDB0000259 | Serotonin | NA | 22.46 | 0.99 (0.86-1.13) | 0.86 | >0.99 | 0.99 (0.83-1.18) | 0.89 | >0.99 |
| HMDB0010370* | CE(18:3) | Cholesteryl esters | 5.65 | 0.94 (0.83-1.06) | 0.32 | >0.99 | 0.99 (0.85-1.15) | 0.90 | >0.99 |
| HMDB0011745 | N-Acetylmethionine | NA | 25.06 | 1.04 (0.92-1.17) | 0.55 | >0.99 | 1.01 (0.87-1.16) | 0.90 | >0.99 |
| HMDB0004956 | Cer(d18:1/24:0) | Ceramides | 8.48 | 1.04 (0.92-1.17) | 0.56 | >0.99 | 0.99 (0.86-1.15) | 0.90 | >0.99 |
| HMDB0008991* | PE(36:0) | Phosphatidylethanolamines | 9.34 | 1.04 (0.91-1.20) | 0.57 | >0.99 | 1.01 (0.84-1.22) | 0.90 | >0.99 |
| HMDB0010405 | LPC(24:0) | Lysophosphatidylcholines | 10.56 | 1.01 (0.90-1.14) | 0.89 | >0.99 | 1.01 (0.87-1.18) | 0.90 | >0.99 |
| HMDB0005923 | N4-Acetylcytidine | Nucleosides, nucleotides, and analogues | 14.84 | 0.91 (0.81-1.03) | 0.14 | >0.99 | 0.99 (0.85-1.16) | 0.91 | >0.99 |
| HMDB0007883* | PC(34:4) | Phosphatidylcholines | 8.41 | 1.05 (0.93-1.19) | 0.41 | >0.99 | 1.01 (0.87-1.17) | 0.91 | >0.99 |
| HMDB0011239* | PC(P-34:0)/PC(O-34:1)_B | Phosphatidylcholine plasmalogens | 13.56 | 1.04 (0.92-1.17) | 0.57 | >0.99 | 1.01 (0.87-1.17) | 0.91 | >0.99 |
| HMDB0011210* | PC(P-34:1)/PC(O-34:2) | Phosphatidylcholine plasmalogens | 8.73 | 1.03 (0.92-1.16) | 0.62 | >0.99 | 1.01 (0.87-1.17) | 0.91 | >0.99 |
| HMDB0007983* | PC(36:4)_A | Phosphatidylcholines | 7.64 | 1.03 (0.91-1.16) | 0.68 | >0.99 | 0.99 (0.85-1.15) | 0.91 | >0.99 |
| HMDB0008925* | PE(34:0) | Phosphatidylethanolamines | 8.57 | 1.02 (0.91-1.15) | 0.73 | >0.99 | 1.01 (0.88-1.16) | 0.91 | >0.99 |
| HMDB0000192 | Cystine | NA | 38.41 | 0.96 (0.84-1.09) | 0.50 | >0.99 | 1.01 (0.86-1.18) | 0.93 | >0.99 |
| HMDB0000206 | N6-Acetyllysine | Organic acids and derivatives | 13.67 | 0.96 (0.86-1.08) | 0.52 | >0.99 | 0.99 (0.86-1.14) | 0.93 | >0.99 |
| NA | Xanthopterin | NA | 26.65 | 0.98 (0.87-1.11) | 0.77 | >0.99 | 1.01 (0.87-1.17) | 0.93 | >0.99 |
| HMDB0013326* | CAR(12:1) | Carnitines | 12.85 | 0.98 (0.87-1.12) | 0.79 | >0.99 | 0.99 (0.86-1.15) | 0.93 | >0.99 |
| HMDB0011442* | PE(P-36:3)/PE(O-36:4) | Phosphatidylethanolamine plasmalogens | 10.68 | 1.01 (0.90-1.13) | 0.89 | >0.99 | 1.01 (0.87-1.16) | 0.93 | >0.99 |
| HMDB0000092 | Dimethylglycine | Carboxylic acids and derivatives | 11.20 | 0.94 (0.83-1.06) | 0.33 | >0.99 | 0.99 (0.86-1.15) | 0.94 | >0.99 |
| HMDB0004620 | N-Acetylarginine | Organic acids and derivatives | 15.22 | 0.98 (0.87-1.10) | 0.70 | >0.99 | 1.01 (0.88-1.15) | 0.94 | >0.99 |
| HMDB0000696 | Methionine | Organic acids and derivatives | 12.19 | 1.01 (0.89-1.14) | 0.88 | >0.99 | 0.99 (0.86-1.15) | 0.94 | >0.99 |
| HMDB0043058* | TG(53:3) | Triglycerides | 7.31 | 1.05 (0.93-1.18) | 0.41 | >0.99 | 1.01 (0.87-1.17) | 0.95 | >0.99 |
| HMDB0000301 | Urocanic acid | NA | 119.02 | 1.00 (0.89-1.13) | 0.94 | >0.99 | 1.00 (0.86-1.15) | 0.95 | >0.99 |
| HMDB0004952 | Cer(d18:1/22:0) | Ceramides | 9.32 | 1.08 (0.96-1.22) | 0.21 | >0.99 | 1.00 (0.86-1.15) | 0.96 | >0.99 |
| HMDB0000158 | Tyrosine | Carboxylic acids and derivatives | 14.63 | 0.99 (0.88-1.11) | 0.84 | >0.99 | 1.00 (0.87-1.16) | 0.96 | >0.99 |
| HMDB0011211* | PC(P-34:2)/PC(O-34:3) | Phosphatidylcholine plasmalogens | 8.28 | 1.00 (0.89-1.12) | 0.98 | >0.99 | 1.00 (0.86-1.15) | 0.96 | >0.99 |
| HMDB0010517* | TG(52:7) | Triglycerides | 12.56 | 1.03 (0.91-1.16) | 0.68 | >0.99 | 1.00 (0.86-1.15) | 0.97 | >0.99 |
| HMDB0002000 | Myristoleic acid | NA | 20.92 | 0.98 (0.87-1.11) | 0.79 | >0.99 | 1.00 (0.85-1.16) | 0.97 | >0.99 |
| HMDB0001991 | 7-Methylxanthine | Organoheterocyclic compounds | 24.44 | 1.00 (0.89-1.12) | 0.94 | >0.99 | 1.00 (0.87-1.15) | 0.97 | >0.99 |
| HMDB0011244* | PC(P-36:2)/PC(O-36:3) | Phosphatidylcholine plasmalogens | 8.42 | 1.02 (0.91-1.14) | 0.74 | >0.99 | 1.00 (0.86-1.15) | 0.98 | >0.99 |
| HMDB0000688 | CAR(5:0) | Carnitines | 21.09 | 0.94 (0.83-1.06) | 0.34 | >0.99 | 1.00 (0.86-1.16) | 0.99 | >0.99 |
| HMDB0012356* | PS(34:0) | Phosphatidylserines | 9.01 | 0.97 (0.86-1.09) | 0.59 | >0.99 | 1.00 (0.87-1.16) | 0.99 | >0.99 |
| HMDB0029416 | Targinine | Organic acids and derivatives | 17.32 | 1.00 (0.87-1.15) | 0.97 | >0.99 | 1.00 (0.83-1.20) | 0.99 | >0.99 |
| HMDB0011131 | MG(18:0) | NA | 37.41 | 1.11 (0.81-1.52) | 0.53 | >0.99 | NA (NA-NA) | NA | NA |
| HMDB0004231 | Pantothenol | NA | 126.90 | 0.79 (0.61-1.02) | 0.07 | >0.99 | NA (NA-NA) | NA | NA |
| HMDB0000965 | Hypotaurine | NA | 29.26 | 0.83 (0.66-1.06) | 0.14 | >0.99 | NA (NA-NA) | NA | NA |
| HMDB0012252 | Linoleoyl-EA | NA | 43.76 | 0.83 (0.63-1.08) | 0.16 | >0.99 | NA (NA-NA) | NA | NA |
| HMDB0011565* | MG(16:1) | NA | 20.65 | 1.20 (0.92-1.57) | 0.17 | >0.99 | NA (NA-NA) | NA | NA |
| HMDB0011241* | PC(P-36:0)/PC(O-36:1) | Phosphatidylcholine plasmalogens | 9.53 | 1.16 (0.92-1.46) | 0.21 | >0.99 | NA (NA-NA) | NA | NA |
| HMDB0006469* | CAR(18:2) | Carnitines | 29.67 | 0.87 (0.68-1.11) | 0.25 | >0.99 | NA (NA-NA) | NA | NA |
| HMDB0001855 | 5-Hydroxytryptophol | Organoheterocyclic compounds | 39.53 | 1.15 (0.90-1.47) | 0.27 | >0.99 | NA (NA-NA) | NA | NA |
| HMDB0003331 | 1-Methyladenosine | NA | 18.24 | 0.88 (0.69-1.12) | 0.30 | >0.99 | NA (NA-NA) | NA | NA |
| HMDB0005065* | CAR(18:1) | Carnitines | 25.54 | 0.88 (0.70-1.12) | 0.30 | >0.99 | NA (NA-NA) | NA | NA |
| HMDB0000222 | CAR(16:0) | Carnitines | 17.61 | 0.89 (0.70-1.12) | 0.32 | >0.99 | NA (NA-NA) | NA | NA |
| HMDB0007170* | DG(38:4) | Diglycerides | 9.19 | 1.11 (0.89-1.40) | 0.36 | >0.99 | NA (NA-NA) | NA | NA |
| HMDB0000292 | Xanthine | NA | 45.61 | 0.91 (0.71-1.17) | 0.47 | >0.99 | NA (NA-NA) | NA | NA |
| HMDB0000252* | Sphingosine | NA | 59.06 | 0.90 (0.66-1.21) | 0.48 | >0.99 | NA (NA-NA) | NA | NA |
| HMDB0012103 | SM(d18:1/22:0) | Sphingomyelins | 8.14 | 1.09 (0.86-1.38) | 0.49 | >0.99 | NA (NA-NA) | NA | NA |
| HMDB0001924 | Atenolol | NA | 8.22 | 0.36 (0.02-7.49) | 0.51 | >0.99 | NA (NA-NA) | NA | NA |
| HMDB0008036* | PC(36:0) | Phosphatidylcholines | 8.60 | 1.08 (0.85-1.39) | 0.52 | >0.99 | NA (NA-NA) | NA | NA |
| HMDB0011319* | PC(P-38:5)/PC(O-38:6) | Phosphatidylcholine plasmalogens | 7.10 | 1.07 (0.85-1.35) | 0.56 | >0.99 | NA (NA-NA) | NA | NA |
| HMDB0000848 | CAR(18:0) | Carnitines | 16.32 | 0.94 (0.74-1.18) | 0.58 | >0.99 | NA (NA-NA) | NA | NA |
| HMDB0000195 | Inosine | NA | 62.75 | 1.06 (0.84-1.34) | 0.61 | >0.99 | NA (NA-NA) | NA | NA |
| HMDB0011294* | PC(P-40:6)/PC(O-40:7) | Phosphatidylcholine plasmalogens | 8.04 | 0.94 (0.74-1.19) | 0.62 | >0.99 | NA (NA-NA) | NA | NA |
| HMDB0007158* | DG(36:0) | Diglycerides | 32.27 | 0.95 (0.70-1.30) | 0.75 | >0.99 | NA (NA-NA) | NA | NA |
| HMDB0013122* | LPC(P-18:0)/LPC(O-18:1) | Phosphatidylcholine plasmalogens | 12.69 | 1.04 (0.81-1.35) | 0.75 | >0.99 | NA (NA-NA) | NA | NA |
| HMDB0010316 | p-Acetamidophenylglucuronide | NA | 18.13 | 1.06 (0.72-1.54) | 0.78 | >0.99 | NA (NA-NA) | NA | NA |
| HMDB0001935 | Warfarin | NA | 31.43 | 1.07 (0.66-1.73) | 0.79 | >0.99 | NA (NA-NA) | NA | NA |
| NA | 3-(N-Acetyl-L-cystein-S-yl) acetaminophen | NA | 32.87 | 1.04 (0.78-1.38) | 0.80 | >0.99 | NA (NA-NA) | NA | NA |
| HMDB0014611 | Quinine | Alkaloids and derivatives | 62.32 | 1.04 (0.76-1.42) | 0.81 | >0.99 | NA (NA-NA) | NA | NA |
| HMDB0000517 | Arginine | NA | 17.38 | 1.03 (0.80-1.33) | 0.82 | >0.99 | NA (NA-NA) | NA | NA |
| HMDB0000269 | Sphinganine | NA | 43.42 | 1.03 (0.80-1.32) | 0.84 | >0.99 | NA (NA-NA) | NA | NA |
| HMDB0001411 | Cotinine N-oxide | NA | 39.22 | 0.95 (0.50-1.82) | 0.88 | >0.99 | NA (NA-NA) | NA | NA |
| HMDB0007970* | PC(34:0) | Phosphatidylcholines | 9.19 | 1.00 (0.79-1.27) | 0.97 | >0.99 | NA (NA-NA) | NA | NA |
| HMDB0001850 | Verapamil | NA | 50.06 | NA (NA-NA) | NA | NA | NA (NA-NA) | NA | NA |
| HMDB0015028 | Sulfapyridine | NA | NA | NA (NA-NA) | NA | NA | NA (NA-NA) | NA | NA |

Abbreviations: CI=confidence interval; CV=coefficient of variation; NA=not available; NEF=number of effective tests; OR=odds ratio

* representative HMDB ID

**Model 1**: basic model, adjusting for matching factors only

**Model 5**: age + smoking status + BMI + physical activity + time of day (as matching imperfect) + month of blood draw (season, as matching imperfect) + family history of POAG + SES + race + age at menopause + nitrate intake + caffeine intake + alcohol intake + alternate healthy eating index + caloric intake + hypertension + high cholesterol + diabetes + oral/inhaled steroid use.

### **Supplementary Table S2. Odds ratios (OR) and 95% confidence intervals (CI) of glaucoma for all metabolites in multivariable-adjusted model in the UK Biobank (2238 glaucoma cases, 44723 controls)**

| **METABOLITE** | **Unit** | **Classification** | **Subclassification** | **Odds ratio (95% CI)** | **p.value** | **NEF-p** |
| --- | --- | --- | --- | --- | --- | --- |
| Tyrosine | mmol/l | Amino acids | Aromatic amino acids | 1.11 (1.06-1.16) | <0.001 | <0.05 |
| Glucose | mmol/l | Glycolysis related metabolites | NA | 1.11 (1.06-1.16) | <0.001 | <0.05 |
| Lactate | mmol/l | Glycolysis related metabolites | NA | 0.84 (0.80-0.87) | <0.001 | <0.05 |
| Pyruvate | mmol/l | Glycolysis related metabolites | NA | 0.89 (0.85-0.93) | <0.001 | <0.05 |
| Citrate | mmol/l | Glycolysis related metabolites | NA | 0.90 (0.86-0.94) | <0.001 | <0.05 |
| 3-Hydroxybutyrate | mmol/l | Ketone bodies | NA | 0.90 (0.86-0.94) | <0.001 | <0.05 |
| Acetate | mmol/l | Ketone bodies | NA | 0.91 (0.87-0.95) | <0.001 | <0.05 |
| Glutamine | mmol/l | Amino acids | NA | 1.07 (1.03-1.12) | <0.01 | >0.99 |
| Triglycerides in IDL | mmol/l | Lipoprotein subclasses | IDL (average diameter 28.6 nm) | 1.09 (1.03-1.14) | <0.01 | >0.99 |
| Triglycerides in Large LDL | mmol/l | Lipoprotein subclasses | Large LDL (average diameter 25.5 nm) | 1.09 (1.03-1.15) | <0.01 | >0.99 |
| Triglycerides in LDL | mmol/l | Triglycerides | NA | 1.08 (1.03-1.14) | <0.01 | >0.99 |
| Triglycerides in Very Small VLDL | mmol/l | Lipoprotein subclasses | Very small VLDL (average diameter 31.3 nm) | 1.07 (1.02-1.13) | 0.01 | >0.99 |
| Triglycerides in Medium LDL | mmol/l | Lipoprotein subclasses | Medium LDL (average diameter 23 nm) | 1.07 (1.02-1.13) | 0.01 | >0.99 |
| Total Lipids in Very Small VLDL | mmol/l | Lipoprotein subclasses | Very small VLDL (average diameter 31.3 nm) | 1.08 (1.01-1.16) | 0.02 | >0.99 |
| Phospholipids in Very Small VLDL | mmol/l | Lipoprotein subclasses | Very small VLDL (average diameter 31.3 nm) | 1.08 (1.01-1.15) | 0.02 | >0.99 |
| Triglycerides in Very Large HDL | mmol/l | Lipoprotein subclasses | Very large HDL (average diameter 14.3 nm) | 1.06 (1.01-1.11) | 0.03 | >0.99 |
| Triglycerides in Small VLDL | mmol/l | Lipoprotein subclasses | Small VLDL (average diameter 36.8 nm) | 1.05 (1.00-1.11) | 0.04 | >0.99 |
| Valine | mmol/l | Amino acids | Branched-chain amino acids | 1.05 (1.00-1.10) | 0.04 | >0.99 |
| Phenylalanine | mmol/l | Amino acids | Aromatic amino acids | 1.05 (1.00-1.09) | 0.04 | >0.99 |
| Concentration of Very Small VLDL Particles | mmol/l | Lipoprotein subclasses | Very small VLDL (average diameter 31.3 nm) | 1.07 (1.00-1.15) | 0.04 | >0.99 |
| Triglycerides in Small LDL | mmol/l | Lipoprotein subclasses | Small LDL (average diameter 18.7 nm) | 1.05 (1.00-1.11) | 0.04 | >0.99 |
| Free Cholesterol in Very Small VLDL | mmol/l | Lipoprotein subclasses | Very small VLDL (average diameter 31.3 nm) | 1.07 (1.00-1.15) | 0.05 | >0.99 |
| Total Triglycerides | mmol/l | Triglycerides | NA | 1.05 (1.00-1.10) | 0.06 | >0.99 |
| Total Lipids in Lipoprotein Particles | mmol/l | Total lipids | NA | 1.08 (1.00-1.16) | 0.06 | >0.99 |
| Total Concentration of Branched-Chain Amino Acids (Leucine + Isoleucine + Valine) | mmol/l | Amino acids | Branched-chain amino acids | 1.04 (1.00-1.09) | 0.07 | >0.99 |
| Saturated Fatty Acids | mmol/l | Fatty acids | NA | 1.05 (1.00-1.12) | 0.07 | >0.99 |
| Phospholipids in Small LDL | mmol/l | Lipoprotein subclasses | Small LDL (average diameter 18.7 nm) | 1.07 (0.99-1.15) | 0.07 | >0.99 |
| Isoleucine | mmol/l | Amino acids | Branched-chain amino acids | 1.04 (1.00-1.09) | 0.08 | >0.99 |
| Triglycerides in HDL | mmol/l | Triglycerides | NA | 1.04 (0.99-1.09) | 0.09 | >0.99 |
| Triglycerides in VLDL | mmol/l | Triglycerides | NA | 1.04 (0.99-1.10) | 0.10 | >0.99 |
| Concentration of Small VLDL Particles | mmol/l | Lipoprotein subclasses | Small VLDL (average diameter 36.8 nm) | 1.05 (0.99-1.11) | 0.10 | >0.99 |
| Cholesterol in Very Small VLDL | mmol/l | Lipoprotein subclasses | Very small VLDL (average diameter 31.3 nm) | 1.07 (0.99-1.15) | 0.10 | >0.99 |
| Concentration of VLDL Particles | mmol/l | Lipoprotein particle concentrations | NA | 1.05 (0.99-1.12) | 0.11 | >0.99 |
| Total Phospholipids in Lipoprotein Particles | mmol/l | Phospholipids | NA | 1.06 (0.99-1.14) | 0.11 | >0.99 |
| Phosphatidylcholines | mmol/l | Other lipids | NA | 1.05 (0.99-1.12) | 0.12 | >0.99 |
| Triglycerides in Medium VLDL | mmol/l | Lipoprotein subclasses | Medium VLDL (average diameter 44.5 nm) | 1.04 (0.99-1.10) | 0.12 | >0.99 |
| Total Lipids in Small VLDL | mmol/l | Lipoprotein subclasses | Small VLDL (average diameter 36.8 nm) | 1.05 (0.99-1.11) | 0.12 | >0.99 |
| Triglycerides in Small HDL | mmol/l | Lipoprotein subclasses | Small HDL (average diameter 8.7 nm) | 1.04 (0.99-1.09) | 0.12 | >0.99 |
| Triglycerides in Very Large VLDL | mmol/l | Lipoprotein subclasses | Very large VLDL (average diameter 64 nm) | 1.04 (0.99-1.09) | 0.14 | >0.99 |
| Triglycerides in Medium HDL | mmol/l | Lipoprotein subclasses | Medium HDL (average diameter 10.9 nm) | 1.04 (0.99-1.09) | 0.14 | >0.99 |
| Phospholipids in Large VLDL | mmol/l | Lipoprotein subclasses | Large VLDL (average diameter 53.6 nm) | 1.04 (0.99-1.09) | 0.14 | >0.99 |
| Triglycerides in Large VLDL | mmol/l | Lipoprotein subclasses | Large VLDL (average diameter 53.6 nm) | 1.04 (0.99-1.09) | 0.14 | >0.99 |
| Glycoprotein Acetyls | mmol/l | Inflammation | NA | 1.04 (0.99-1.09) | 0.14 | >0.99 |
| Total Lipids in VLDL | mmol/l | Total lipids | NA | 1.04 (0.99-1.10) | 0.15 | >0.99 |
| Phospholipids in VLDL | mmol/l | Phospholipids | NA | 1.04 (0.98-1.10) | 0.16 | >0.99 |
| Triglycerides in Large HDL | mmol/l | Lipoprotein subclasses | Large HDL (average diameter 12.1 nm) | 1.04 (0.99-1.09) | 0.16 | >0.99 |
| Docosahexaenoic Acid | mmol/l | Fatty acids | NA | 1.04 (0.99-1.09) | 0.16 | >0.99 |
| Cholesteryl Esters in Very Small VLDL | mmol/l | Lipoprotein subclasses | Very small VLDL (average diameter 31.3 nm) | 1.06 (0.98-1.14) | 0.16 | >0.99 |
| Total Lipids in Large VLDL | mmol/l | Lipoprotein subclasses | Large VLDL (average diameter 53.6 nm) | 1.04 (0.98-1.09) | 0.17 | >0.99 |
| Concentration of Large VLDL Particles | mmol/l | Lipoprotein subclasses | Very large VLDL (average diameter 64 nm) | 1.04 (0.98-1.09) | 0.17 | >0.99 |
| Concentration of Very Large VLDL Particles | mmol/l | Lipoprotein subclasses | Very large VLDL (average diameter 64 nm) | 1.04 (0.98-1.09) | 0.18 | >0.99 |
| Total Fatty Acids | mmol/l | Fatty acids | NA | 1.04 (0.98-1.10) | 0.19 | >0.99 |
| Leucine | mmol/l | Amino acids | Branched-chain amino acids | 1.03 (0.98-1.08) | 0.19 | >0.99 |
| Concentration of Small LDL Particles | mmol/l | Lipoprotein subclasses | Small LDL (average diameter 18.7 nm) | 1.05 (0.98-1.13) | 0.19 | >0.99 |
| Phospholipids in Very Large VLDL | mmol/l | Lipoprotein subclasses | Very large VLDL (average diameter 64 nm) | 1.03 (0.98-1.09) | 0.20 | >0.99 |
| Total Lipids in Medium VLDL | mmol/l | Lipoprotein subclasses | Medium VLDL (average diameter 44.5 nm) | 1.04 (0.98-1.11) | 0.21 | >0.99 |
| Total Lipids in Very Large VLDL | mmol/l | Lipoprotein subclasses | Very large VLDL (average diameter 64 nm) | 1.03 (0.98-1.09) | 0.21 | >0.99 |
| Omega-3 Fatty Acids | mmol/l | Fatty acids | NA | 1.03 (0.98-1.08) | 0.22 | >0.99 |
| Phospholipids in Small VLDL | mmol/l | Lipoprotein subclasses | Small VLDL (average diameter 36.8 nm) | 1.04 (0.98-1.11) | 0.23 | >0.99 |
| Total Lipids in IDL | mmol/l | Lipoprotein subclasses | IDL (average diameter 28.6 nm) | 1.06 (0.97-1.15) | 0.23 | >0.99 |
| Cholesteryl Esters in Small HDL | mmol/l | Lipoprotein subclasses | Small HDL (average diameter 8.7 nm) | 0.97 (0.93-1.02) | 0.23 | >0.99 |
| Phosphoglycerides | mmol/l | Other lipids | NA | 1.04 (0.98-1.10) | 0.23 | >0.99 |
| Free Cholesterol in VLDL | mmol/l | Free cholesterol | NA | 1.04 (0.98-1.10) | 0.24 | >0.99 |
| Free Cholesterol in Large VLDL | mmol/l | Lipoprotein subclasses | Large VLDL (average diameter 53.6 nm) | 1.03 (0.98-1.09) | 0.24 | >0.99 |
| Phospholipids in IDL | mmol/l | Lipoprotein subclasses | IDL (average diameter 28.6 nm) | 1.05 (0.96-1.15) | 0.25 | >0.99 |
| Phospholipids in Medium VLDL | mmol/l | Lipoprotein subclasses | Medium VLDL (average diameter 44.5 nm) | 1.04 (0.97-1.11) | 0.26 | >0.99 |
| Total Free Cholesterol | mmol/l | Free cholesterol | NA | 1.05 (0.96-1.15) | 0.26 | >0.99 |
| Albumin | g/l | Fluid balance | NA | 0.98 (0.93-1.02) | 0.26 | >0.99 |
| Free Cholesterol in Very Large VLDL | mmol/l | Lipoprotein subclasses | Very large VLDL (average diameter 64 nm) | 1.03 (0.98-1.08) | 0.27 | >0.99 |
| Total Lipids in Small LDL | mmol/l | Lipoprotein subclasses | Small LDL (average diameter 18.7 nm) | 1.04 (0.97-1.12) | 0.28 | >0.99 |
| Free Cholesterol in Very Large HDL | mmol/l | Lipoprotein subclasses | Very large HDL (average diameter 14.3 nm) | 1.03 (0.98-1.08) | 0.29 | >0.99 |
| Creatinine | mmol/l | Fluid balance | NA | 0.97 (0.92-1.02) | 0.29 | >0.99 |
| Remnant Cholesterol (Non-HDL, Non-LDL -Cholesterol) | mmol/l | Cholesterol | NA | 1.05 (0.96-1.14) | 0.29 | >0.99 |
| Apolipoprotein B | g/l | Apolipoproteins | NA | 1.04 (0.96-1.13) | 0.30 | >0.99 |
| Phospholipids in Very Large HDL | mmol/l | Lipoprotein subclasses | Very large HDL (average diameter 14.3 nm) | 1.03 (0.98-1.08) | 0.30 | >0.99 |
| Cholesterol in Large VLDL | mmol/l | Lipoprotein subclasses | Large VLDL (average diameter 53.6 nm) | 1.03 (0.97-1.09) | 0.31 | >0.99 |
| Total Lipids in Very Large HDL | mmol/l | Lipoprotein subclasses | Very large HDL (average diameter 14.3 nm) | 1.03 (0.97-1.08) | 0.33 | >0.99 |
| Cholesteryl Esters in IDL | mmol/l | Lipoprotein subclasses | IDL (average diameter 28.6 nm) | 1.04 (0.96-1.13) | 0.33 | >0.99 |
| Concentration of Large LDL Particles | mmol/l | Lipoprotein subclasses | Large LDL (average diameter 25.5 nm) | 1.04 (0.96-1.12) | 0.34 | >0.99 |
| Concentration of Very Large HDL Particles | mmol/l | Lipoprotein subclasses | Very large HDL (average diameter 14.3 nm) | 1.03 (0.97-1.08) | 0.35 | >0.99 |
| Free Cholesterol in Small HDL | mmol/l | Lipoprotein subclasses | Small HDL (average diameter 8.7 nm) | 1.03 (0.97-1.08) | 0.36 | >0.99 |
| Total Cholines | mmol/l | Other lipids | NA | 1.03 (0.97-1.10) | 0.36 | >0.99 |
| Free Cholesterol in Medium VLDL | mmol/l | Lipoprotein subclasses | Medium VLDL (average diameter 44.5 nm) | 1.03 (0.96-1.11) | 0.38 | >0.99 |
| Free Cholesterol in Small VLDL | mmol/l | Lipoprotein subclasses | Small VLDL (average diameter 36.8 nm) | 1.03 (0.96-1.11) | 0.38 | >0.99 |
| VLDL Cholesterol | mmol/l | Cholesterol | NA | 1.03 (0.96-1.10) | 0.38 | >0.99 |
| Alanine | mmol/l | Amino acids | NA | 0.98 (0.94-1.03) | 0.39 | >0.99 |
| Polyunsaturated Fatty Acids | mmol/l | Fatty acids | NA | 1.03 (0.96-1.10) | 0.40 | >0.99 |
| Concentration of LDL Particles | mmol/l | Lipoprotein particle concentrations | NA | 1.03 (0.96-1.12) | 0.40 | >0.99 |
| Cholesterol in Very Large VLDL | mmol/l | Lipoprotein subclasses | Very large VLDL (average diameter 64 nm) | 1.02 (0.97-1.08) | 0.41 | >0.99 |
| Total Cholesterol Minus HDL-C | mmol/l | Cholesterol | NA | 1.04 (0.95-1.13) | 0.41 | >0.99 |
| Concentration of Medium VLDL Particles | mmol/l | Lipoprotein subclasses | Medium VLDL (average diameter 44.5 nm) | 1.03 (0.96-1.10) | 0.42 | >0.99 |
| Cholesteryl Esters in Large LDL | mmol/l | Lipoprotein subclasses | Large LDL (average diameter 25.5 nm) | 1.03 (0.96-1.12) | 0.42 | >0.99 |
| Cholesterol in Small VLDL | mmol/l | Lipoprotein subclasses | Small VLDL (average diameter 36.8 nm) | 1.03 (0.96-1.10) | 0.43 | >0.99 |
| Cholesteryl Esters in Large VLDL | mmol/l | Lipoprotein subclasses | Large VLDL (average diameter 53.6 nm) | 1.02 (0.97-1.08) | 0.43 | >0.99 |
| Cholesteryl Esters in Small VLDL | mmol/l | Lipoprotein subclasses | Small VLDL (average diameter 36.8 nm) | 1.03 (0.96-1.09) | 0.44 | >0.99 |
| Concentration of IDL Particles | mmol/l | Lipoprotein subclasses | IDL (average diameter 28.6 nm) | 1.03 (0.95-1.12) | 0.45 | >0.99 |
| Total Lipids in LDL | mmol/l | Total lipids | NA | 1.03 (0.95-1.11) | 0.46 | >0.99 |
| Free Cholesterol in HDL | mmol/l | Free cholesterol | NA | 1.02 (0.97-1.08) | 0.46 | >0.99 |
| Histidine | mmol/l | Amino acids | NA | 0.98 (0.94-1.03) | 0.47 | >0.99 |
| Total Cholesterol | mmol/l | Cholesterol | NA | 1.03 (0.95-1.13) | 0.47 | >0.99 |
| Free Cholesterol in Large HDL | mmol/l | Lipoprotein subclasses | Large HDL (average diameter 12.1 nm) | 1.02 (0.97-1.08) | 0.47 | >0.99 |
| Total Lipids in Large LDL | mmol/l | Lipoprotein subclasses | Large LDL (average diameter 25.5 nm) | 1.03 (0.95-1.12) | 0.48 | >0.99 |
| Cholesterol in Small HDL | mmol/l | Lipoprotein subclasses | Small HDL (average diameter 8.7 nm) | 0.98 (0.94-1.03) | 0.48 | >0.99 |
| Cholesterol in IDL | mmol/l | Lipoprotein subclasses | IDL (average diameter 28.6 nm) | 1.03 (0.95-1.12) | 0.48 | >0.99 |
| Cholesterol in Very Large HDL | mmol/l | Lipoprotein subclasses | Very large HDL (average diameter 14.3 nm) | 1.02 (0.97-1.07) | 0.49 | >0.99 |
| Cholesteryl Esters in LDL | mmol/l | Cholesteryl esters | NA | 1.03 (0.95-1.11) | 0.49 | >0.99 |
| Phospholipids in Medium LDL | mmol/l | Lipoprotein subclasses | Medium LDL (average diameter 23 nm) | 1.03 (0.96-1.10) | 0.49 | >0.99 |
| Acetone | mmol/l | Ketone bodies | NA | 0.99 (0.94-1.03) | 0.50 | >0.99 |
| Concentration of Small HDL Particles | mmol/l | Lipoprotein subclasses | Small HDL (average diameter 8.7 nm) | 0.98 (0.94-1.03) | 0.54 | >0.99 |
| Monounsaturated Fatty Acids | mmol/l | Fatty acids | NA | 1.02 (0.96-1.07) | 0.54 | >0.99 |
| Phospholipids in LDL | mmol/l | Phospholipids | NA | 1.02 (0.95-1.11) | 0.55 | >0.99 |
| Total Lipids in Medium LDL | mmol/l | Lipoprotein subclasses | Medium LDL (average diameter 23 nm) | 1.02 (0.95-1.10) | 0.55 | >0.99 |
| Concentration of Chylomicrons and Extremely Large VLDL Particles | mmol/l | Lipoprotein subclasses | Chylomicrons and extremely large VLDL (particle diameters from 75 nm upwards) | 1.01 (0.97-1.07) | 0.56 | >0.99 |
| Cholesteryl Esters in Very Large HDL | mmol/l | Lipoprotein subclasses | Very large HDL (average diameter 14.3 nm) | 1.02 (0.96-1.07) | 0.56 | >0.99 |
| Degree of Unsaturation |  | Fatty acids | NA | 0.99 (0.94-1.03) | 0.57 | >0.99 |
| Total Esterified Cholesterol | mmol/l | Cholesteryl esters | NA | 1.02 (0.94-1.11) | 0.58 | >0.99 |
| Cholesteryl Esters in VLDL | mmol/l | Cholesteryl esters | NA | 1.02 (0.95-1.10) | 0.59 | >0.99 |
| Phospholipids in Large HDL | mmol/l | Lipoprotein subclasses | Large HDL (average diameter 12.1 nm) | 1.01 (0.96-1.07) | 0.61 | >0.99 |
| Phospholipids in HDL | mmol/l | Phospholipids | NA | 1.01 (0.96-1.07) | 0.61 | >0.99 |
| Triglycerides in Chylomicrons and Extremely Large VLDL | mmol/l | Lipoprotein subclasses | Chylomicrons and extremely large VLDL (particle diameters from 75 nm upwards) | 1.01 (0.97-1.06) | 0.61 | >0.99 |
| Cholesterol in Large LDL | mmol/l | Lipoprotein subclasses | Large LDL (average diameter 25.5 nm) | 1.02 (0.94-1.10) | 0.62 | >0.99 |
| Cholesteryl Esters in Medium HDL | mmol/l | Lipoprotein subclasses | Medium HDL (average diameter 10.9 nm) | 0.99 (0.94-1.04) | 0.63 | >0.99 |
| Average Diameter for HDL Particles | nm | Lipoprotein particle sizes | NA | 1.01 (0.96-1.07) | 0.64 | >0.99 |
| Cholesteryl Esters in Very Large VLDL | mmol/l | Lipoprotein subclasses | Very large VLDL (average diameter 64 nm) | 1.01 (0.96-1.07) | 0.64 | >0.99 |
| Average Diameter for VLDL Particles | nm | Lipoprotein particle sizes | NA | 1.01 (0.96-1.06) | 0.65 | >0.99 |
| LDL Cholesterol | mmol/l | Cholesterol | NA | 1.02 (0.94-1.10) | 0.66 | >0.99 |
| Phospholipids in Medium HDL | mmol/l | Lipoprotein subclasses | Medium HDL (average diameter 10.9 nm) | 1.01 (0.96-1.06) | 0.67 | >0.99 |
| Cholesteryl Esters in Medium LDL | mmol/l | Lipoprotein subclasses | Medium LDL (average diameter 23 nm) | 1.02 (0.95-1.09) | 0.68 | >0.99 |
| Cholesteryl Esters in HDL | mmol/l | Cholesteryl esters | NA | 0.99 (0.94-1.04) | 0.69 | >0.99 |
| Cholesterol in Small LDL | mmol/l | Lipoprotein subclasses | Small LDL (average diameter 18.7 nm) | 1.02 (0.94-1.09) | 0.69 | >0.99 |
| Total Lipids in Large HDL | mmol/l | Lipoprotein subclasses | Large HDL (average diameter 12.1 nm) | 1.01 (0.96-1.07) | 0.69 | >0.99 |
| Free Cholesterol in Chylomicrons and Extremely Large VLDL | mmol/l | Lipoprotein subclasses | Chylomicrons and extremely large VLDL (particle diameters from 75 nm upwards) | 1.01 (0.96-1.06) | 0.70 | >0.99 |
| Omega-6 Fatty Acids | mmol/l | Fatty acids | NA | 1.01 (0.95-1.08) | 0.70 | >0.99 |
| Sphingomyelins | mmol/l | Other lipids | NA | 1.01 (0.95-1.09) | 0.70 | >0.99 |
| Cholesteryl Esters in Small LDL | mmol/l | Lipoprotein subclasses | Small LDL (average diameter 18.7 nm) | 1.01 (0.94-1.09) | 0.70 | >0.99 |
| Phospholipids in Chylomicrons and Extremely Large VLDL | mmol/l | Lipoprotein subclasses | Chylomicrons and extremely large VLDL (particle diameters from 75 nm upwards) | 1.01 (0.96-1.06) | 0.71 | >0.99 |
| Glycine | mmol/l | Amino acids | NA | 1.01 (0.96-1.06) | 0.72 | >0.99 |
| Total Lipids in Chylomicrons and Extremely Large VLDL | mmol/l | Lipoprotein subclasses | Chylomicrons and extremely large VLDL (particle diameters from 75 nm upwards) | 1.01 (0.96-1.06) | 0.72 | >0.99 |
| Free Cholesterol in Medium HDL | mmol/l | Lipoprotein subclasses | Medium HDL (average diameter 10.9 nm) | 1.01 (0.96-1.06) | 0.73 | >0.99 |
| Linoleic Acid | mmol/l | Fatty acids | NA | 1.01 (0.95-1.08) | 0.74 | >0.99 |
| Cholesteryl Esters in Medium VLDL | mmol/l | Lipoprotein subclasses | Medium VLDL (average diameter 44.5 nm) | 0.99 (0.92-1.06) | 0.74 | >0.99 |
| Free Cholesterol in Large LDL | mmol/l | Lipoprotein subclasses | Large LDL (average diameter 25.5 nm) | 0.99 (0.91-1.07) | 0.74 | >0.99 |
| Cholesterol in Medium HDL | mmol/l | Lipoprotein subclasses | Medium HDL (average diameter 10.9 nm) | 0.99 (0.94-1.04) | 0.75 | >0.99 |
| Free Cholesterol in Small LDL | mmol/l | Lipoprotein subclasses | Small LDL (average diameter 18.7 nm) | 1.01 (0.95-1.08) | 0.75 | >0.99 |
| Cholesterol in Medium LDL | mmol/l | Lipoprotein subclasses | Medium LDL (average diameter 23 nm) | 1.01 (0.94-1.09) | 0.76 | >0.99 |
| Total Lipids in HDL | mmol/l | Total lipids | NA | 1.01 (0.96-1.06) | 0.76 | >0.99 |
| Phospholipids in Small HDL | mmol/l | Lipoprotein subclasses | Small HDL (average diameter 8.7 nm) | 1.01 (0.96-1.06) | 0.78 | >0.99 |
| Concentration of HDL Particles | mmol/l | Lipoprotein particle concentrations | NA | 0.99 (0.94-1.05) | 0.80 | >0.99 |
| Concentration of Large HDL Particles | mmol/l | Lipoprotein subclasses | Large HDL (average diameter 12.1 nm) | 1.01 (0.95-1.06) | 0.83 | >0.99 |
| Cholesterol in Medium VLDL | mmol/l | Lipoprotein subclasses | Medium VLDL (average diameter 44.5 nm) | 1.01 (0.93-1.09) | 0.83 | >0.99 |
| Phospholipids in Large LDL | mmol/l | Lipoprotein subclasses | Large LDL (average diameter 25.5 nm) | 1.01 (0.93-1.09) | 0.84 | >0.99 |
| Free Cholesterol in LDL | mmol/l | Free cholesterol | NA | 0.99 (0.92-1.07) | 0.85 | >0.99 |
| HDL Cholesterol | mmol/l | Cholesterol | NA | 1.00 (0.94-1.05) | 0.87 | >0.99 |
| Total Concentration of Lipoprotein Particles | mmol/l | Lipoprotein particle concentrations | NA | 1.00 (0.94-1.05) | 0.89 | >0.99 |
| Total Lipids in Medium HDL | mmol/l | Lipoprotein subclasses | Medium HDL (average diameter 10.9 nm) | 1.00 (0.95-1.06) | 0.89 | >0.99 |
| Cholesterol in Large HDL | mmol/l | Lipoprotein subclasses | Large HDL (average diameter 12.1 nm) | 1.00 (0.95-1.06) | 0.89 | >0.99 |
| Average Diameter for LDL Particles | nm | Lipoprotein particle sizes | NA | 1.00 (0.96-1.05) | 0.89 | >0.99 |
| Concentration of Medium LDL Particles | mmol/l | Lipoprotein subclasses | Medium LDL (average diameter 23 nm) | 1.00 (0.94-1.08) | 0.91 | >0.99 |
| Cholesterol in Chylomicrons and Extremely Large VLDL | mmol/l | Lipoprotein subclasses | Chylomicrons and extremely large VLDL (particle diameters from 75 nm upwards) | 1.00 (0.95-1.05) | 0.92 | >0.99 |
| Cholesteryl Esters in Chylomicrons and Extremely Large VLDL | mmol/l | Lipoprotein subclasses | Chylomicrons and extremely large VLDL (particle diameters from 75 nm upwards) | 1.00 (0.95-1.05) | 0.93 | >0.99 |
| Concentration of Medium HDL Particles | mmol/l | Lipoprotein subclasses | Medium HDL (average diameter 10.9 nm) | 1.00 (0.95-1.05) | 0.94 | >0.99 |
| Total Lipids in Small HDL | mmol/l | Lipoprotein subclasses | Small HDL (average diameter 8.7 nm) | 1.00 (0.95-1.05) | 0.94 | >0.99 |
| Clinical LDL Cholesterol | mmol/l | Cholesterol | NA | 1.00 (0.93-1.08) | 0.95 | >0.99 |
| Free Cholesterol in IDL | mmol/l | Lipoprotein subclasses | IDL (average diameter 28.6 nm) | 1.00 (0.92-1.08) | 0.96 | >0.99 |
| Free Cholesterol in Medium LDL | mmol/l | Lipoprotein subclasses | Medium LDL (average diameter 23 nm) | 1.00 (0.93-1.07) | 0.96 | >0.99 |
| Apolipoprotein A1 | g/l | Apolipoproteins | NA | 1.00 (0.95-1.05) | 0.96 | >0.99 |
| Cholesteryl Esters in Large HDL | mmol/l | Lipoprotein subclasses | Large HDL (average diameter 12.1 nm) | 1.00 (0.95-1.05) | 0.97 | >0.99 |
| Acetoacetate | mmol/l | Ketone bodies | NA | 1.00 (0.96-1.04) | 0.99 | >0.99 |

Abbreviations: CI=confidence interval; OR=odds ratio

Multiple logistic regression model includes age, sex, smoking status, physical activity, BMI, ethnicity, spherical equivalent, coffee consumption, tea consumption, alcohol intake, systolic blood pressure, cholesterol level, diabetes, coronary artery disease, and statin use.
